# Decoding Humoral Immunity During Acute MPXV Infection via Comprehensive Serological Analysis and Antigen-agnostic Monoclonal Antibody Profiling

**DOI:** 10.64898/2026.08.21.26360138

**Authors:** Yifan Zhang, Jingxian Fan, Jing Wang, Ning Jiang, Yanmin Wan, Lu Meng, Weiqiang Qi, Xiaoyang Cheng, Kexin Luo, Tengfei Zhang, Ronghui Li, Haili Chen, Ranjie Zhao, Yanqin Ren, Wenhong Zhang, Zhaoqin Zhu

## Abstract

Dissecting the complexity of antibody responses in orthopoxvirus (OPXV) infected individuals is essential for elucidating protective mechanisms and identifying candidate protective immunogens. Here, we profiled the acute humoral response in 51 mpox cases, showing distinct IgG trajectories among multiple antigens alongside the rise of plasma neutralizing activities to plateau within 6 weeks after symptom onset. Utilizing a single-cell transcriptomic and BCR sequencing based antigen-agnostic mAb isolation workflow, we further generated monoclonal antibodies (mAbs) from 254 expanded peripheral B cell clones of 3 patients. We discerned 97 specific mAbs recognizing at least 12 different OPXV proteins via integrated screening approaches, which comprised neutralizing antibodies binding unconventional viral targets and antibodies exhibiting extraordinary in vitro and in vivo anti-OPXV effects. The number of OPXV-specific mAbs recovered per donor reflected the percentage of expanded clones among circulating B cells. More interestingly, we demonstrated that the inferred unmutated common ancestors (UCAs) of neutralizing antibody clones did not necessarily react with OPXV, implying that OPXV neutralizing antibodies might frequently originate from B cells previously activated by unknown antigens. Our work establishes an efficient workflow for antigen-agnostic isolation of pathogen specific mAbs and reveals previously unclarified features of antibody responses induced by acute MPXV infection.

## Introduction

MPXV is a member of the OPXV genus, which includes four other human pathogenic species, variola virus (VARV), cowpox virus, camelpox virus and vaccinia virus (VACV) [1]. Given that OPXVs share high antigenic similarity, smallpox vaccination campaign not only eradicated smallpox but also established herd immunity against other OPXVs, which helped to curb the outbreak and spread of MPXV worldwide [2, 3]. However, recent mpox outbreaks have underscored the persistent public health threat posed by OPXVs [4–6], particularly as population-level immunity has waned [7, 8].

Although both antibody and T cell responses were suggested to play a role in protection against OPXV infection [9], neutralizing antibody response is thought to be a necessary and sufficient factor [10, 11]. Multiple proteins encoded by OPXVs have been identified as neutralizing determinants, including VACV A27, D8, H3, A28, F9, A16/G9, A26, L1, A13, A17 on the surface of intracellular mature virion (IMV) and A33, B5 on the surface of extracellular enveloped virion (EEV) [12–19]. A fundamental feature of antibody responses upon smallpox vaccination or OPXV infection is that antibodies to multiple targets together constitute the redundant protective humoral responses [20], but the serological specificities can vary from individual to individual [20, 21] and it seems that no single target specificity is indispensable.

Isolating neutralizing antibodies from OPXV infected individuals and characterizing their antigen specificities represents a crucial way to deconvolute the complicated protective humoral immunity to this virus [22]. Several previous studies successfully isolated monoclonal neutralizing antibodies from individuals inoculated with a smallpox vaccine or infected by MPXV using antigen-defined methods [15–19, 23, 24], but these methods were incapable of identifying antibodies targeting distinct or unknown antigens. Two studies used antigen-agnostic antibody recovery approaches to capture the natural diversity of OPXV specific antibody responses in humans [25, 26], of which only one study characterized the neutralizing activities for the isolated mAbs and suggested that VACV D8, L1, B5, A33, A27, H3 are the major neutralizing determinants [26]. These studies have improved our knowledge about the complexity of antibody responses against OPXVs, but are still far from fully resolving it.

In this study, we comprehensively investigated the specific humoral immunity during acute MPXV infection by integrating plasma serology, single-cell transcriptomics, paired BCR sequencing, and B-cell clonal expansion guided mAb recovery and functional characterization. This strategy yielded a diverse panel of OPXV-reactive mAbs against at least 12 viral proteins, including potent neutralizing antibodies against established IMV and EEV targets as well as antibodies with unexpected antigen specificities. Notably, we identified a B6-reactive mAb, which recognizes a linear epitope (S220-L234) within the membrane-proximal SCR4 domain and confers robust protection across both respiratory and systemic VACV challenge mouse models. The B-cell clonal expansion-guided antibody recovery strategy also enabled analysis of OPXV-reactive antibody lineages, and examination of representative clones showed that some inferred UCAs retained OPXV reactivity, whereas others lacked detectable binding and neutralizing activity, raising the possibility that a subset of OPXV-neutralizing antibody lineages originated from B cells primed by unknown antigens before MPXV infection. Together, these findings establish expanded circulating B-cell clonotypes as an effective indicator for antigen-agnostic OPXV antibody discovery and reveal previously unrecognized features of acute antibody responses to MPXV infection.

## Results

### Neutralizing antibodies elicited by natural MPXV infection increased gradually within 6 weeks since symptom onset, while the kinetics of binding antibody responses varied substantially against different viral targets

51 laboratory-confirmed, hospitalized mpox patients were included in this study. The median age was 32 years (IQR, 28-37 years), most patients (46, 90.2%) were men who have sex with men, 20 (39.2%) patients were HIV-positive, and 9 (17.6%) patients presented concurrent non-HIV sexually transmitted infections (Supplementary Table 1). These patients had neither a history of smallpox vaccination nor prior OPXV infection, and did not receive anti-orthopoxvirus treatment. MPXV DNA was detectable in plasma of 24 (47.1%) patients at the time of sampling (Supplementary Table 1). In a previous work, we retrospectively looked into the plasma binding antibody responses to whole virus lysate (WVL) of Tiantan vaccinia virus and neutralizing antibody responses against the IMV of Tiantan vaccinia virus, which revealed no significant differences between HIV-positive and HIV-negative patients [27].

Building on this preliminary observation, this study further dissected the specific antibody responses ensuing from MPXV infection. We divided these patients into 6 groups according to their peripheral blood sampling time points (0-2, 3-4, 5-6, 7-8, 9-14 and 15-38 days since symptom onset) and quantified plasma neutralizing activities against both the IMV and EEV forms of Tiantan vaccinia virus for each sample (Supplementary Figure 1). We found that the areas under both inhibition curves (AUCs) increased over time within 7-8 days post symptom onset and remained stable till 15-38 days (Figure 1A). Neither the WVL binding IgG nor the binding IgM responses fully synchronized with the plasma neutralizing activities (Figure 1B). The WVL binding IgG responses continued to increase at the last sampling time window, while the WVL binding IgM responses declined at 15-38 days post symptom onset (Figure 1B).

**Figure 1.**
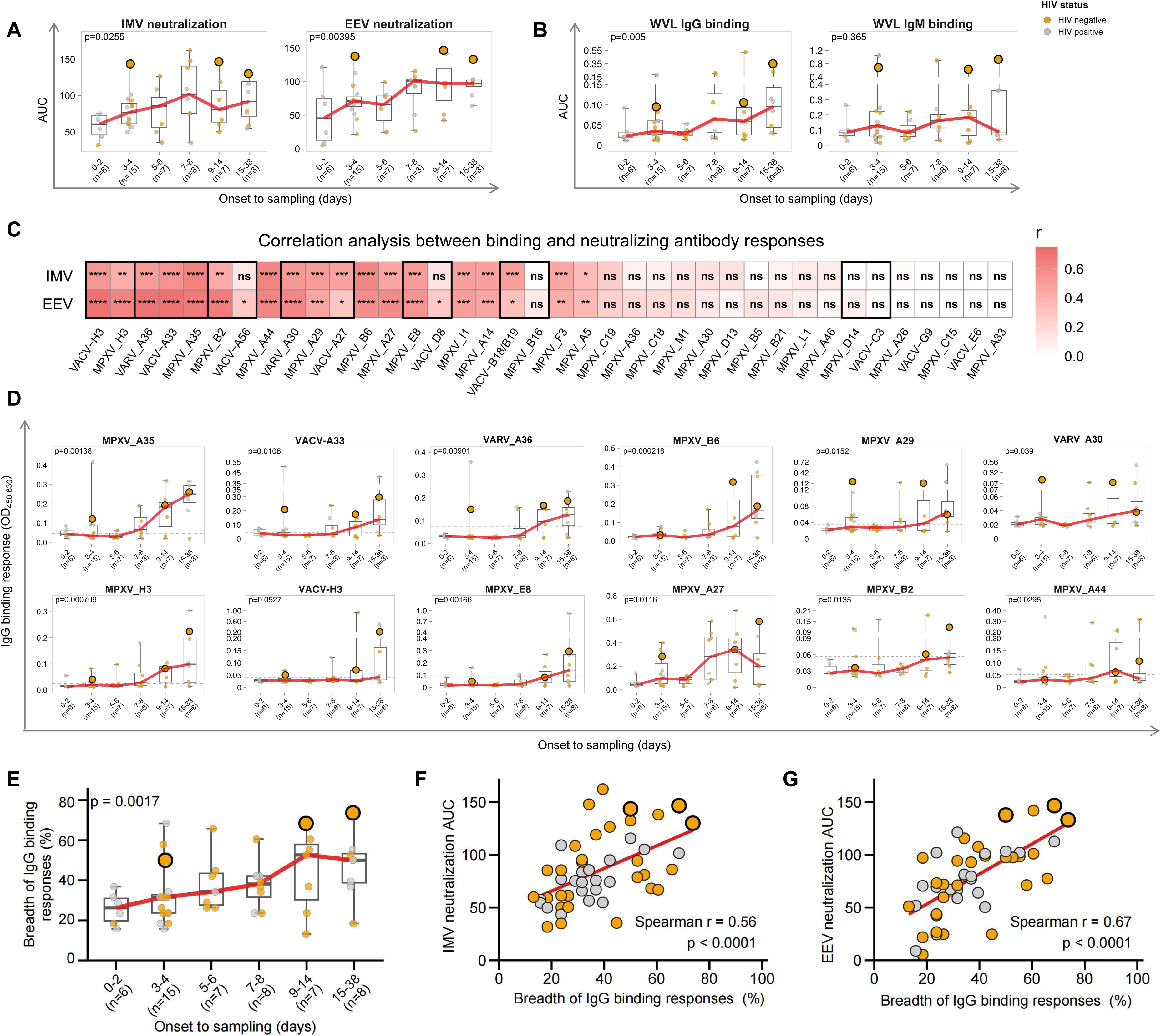
Characterization of serological responses induced by acute MPXV infection. **(A)** Neutralizing activities of mpox patients’ plasma against the IMV and EEV forms of Tiantan vaccinia virus. Neutralizing activities are displayed as the area under the plaque reduction against plasma dilution curve. **(B)** The time course analyses of WVL specific IgG and IgM responses. Binding activities are expressed as the area under the absorbance (OD) against plasma dilution curve. **(C)** Spearman correlation analyses between the AUCs of neutralizing antibody responses and the OD values of antigen specific IgG responses. The Spearman correlation coefficient (r) is indicated by color intensity, and statistical significance is shown in each cell. Black borders highlight groups of orthologous proteins from different OPXV species. ****, p < 0.0001; ***, p < 0.001; **, p < 0.01; *, p < 0.05; ns, p ≥ 0.05. **(D)** The kinetics of antigen specific IgG binding responses that strongly (r ≥ 0.5) correlate with the plasma neutralizing activities. Dashed lines indicate the cutoff values defined as twice the OD_450-630_ of a negative control sample. (**E**) The time course analysis for the breadth of binding IgG responses. (**F**, **G**) Correlations between the breadth of binding IgG responses and IMV (**F**) or EEV (**G**) neutralizing activity. Data in (**A**), (**B**), (**D**), and (**E**) are presented as medians with IQRs. Each circle represents one sample, and fill color indicates HIV status. The enlarged circles indicate the three samples selected for subsequent single-cell analysis. P values in (**A**), (**B**), (**D**), and (**E**) were calculated using the two-sided Jonckheere-Terpstra test for ordered trends across time windows of sample collection.

MPXV encodes 181 proteins [28] and previous studies suggested that serological responses of mpox patients varied across a few putative neutralizing targets [29–31]. In this study, we expanded the measurement of binding IgG responses to 27 MPXV proteins, 9 VACV proteins and 2 VARV proteins, which are summarized into four groups according to intra-virion protein localization (Supplementary Table 2). Our data show that the binding IgG responses against 20 proteins positively correlate with the plasma neutralizing activities towards the IMV or EEV form of Tiantan vaccinia virus, including 6 EEV membrane proteins (MPXV A35, B2 and B6, VACV A33 and A56, and VARV A36), 9 IMV membrane proteins (MPXV A14, A27, A29, E8 and H3, VACV A27, H3 and D8, and VARV A30) and 5 non-membrane associated viral proteins (MPXV A5, F3, I1 and A44, and VACV B18/B19) (Figure 1C). Specific IgG responses towards 12 proteins correlated relatively strongly (Spearman r ≥ 0.5) with plasma neutralizing activities (Figure 1D), while the correlations between IgG responses against the other 8 proteins and plasma neutralizing activities were weak (Spearman r < 0.5) (Supplementary Figure 2A). The time courses of IgG responses against proteins of both categories were heterogeneous, most of which showed a tendency of increasing over time, but only the binding IgG responses against MPXV A35, H3, A27, A14, I1 and VACV D8 showed median OD values higher than their corresponding cutoff values at 7-8 days (Figure 1D and Supplementary Figure 2A), when the average plasma neutralizing responses increased to plateau levels (Figure 1A). IgG responses to the remaining 18 viral antigens listed in Supplementary Table 2 showed no significant correlation with either IMV or EEV neutralizing activity (Figure 1C) and specific IgG responses against these proteins did not show consistent tendencies of increasing over time (Supplementary Figure 2B). We next asked whether the breadth of antigen recognition could reflect plasma neutralizing activity. Herein, the breadth of the IgG binding response was defined as the percentage of tested antigens that exhibited binding signals above their corresponding cutoff values. We found that binding IgG breadth increased from a median of 26.3% at 0-2 days to 52.6% at 9-14 days and maintained 50.0% at 15-38 days (Figure 1E). Moreover, the breadth of antigen recognition positively correlated with both IMV and EEV neutralizing activities (Figures 1F and 1G), indicating that stronger plasma neutralization is associated with broader IgG responses.

### Single-cell transcriptomic and BCR repertoire sequencing revealed dynamic clonal expansion of circulating B cells driven by MPXV infection

The above results reveal that the serological neutralizing activities rise even faster than the binding IgG responses against most viral proteins detected in this study (Figures 1A, 1D and Supplementary Figure 2). We reason that neutralizing antibody responses elicited by MPXV infections might resemble the feature of neutralizing antibody responses elicited by smallpox vaccine, which are mediated by a collection of highly redundant specific antibodies to multiple viral proteins and can vary from individual to individual [20]. To gain a deeper insight into the early humoral responses upon MPXV infection, we performed single-cell transcriptomic and B-cell receptor (BCR) sequencing for PBMC samples of three HIV and STI negative patients selected based on serological measurements and sampling time point (Figure 2A). Samples YY_D3, LXD_D10, and YHB_D18 were collected at 3, 10, and 18 days post-symptom onset, respectively. All three samples exhibited appreciable neutralizing and binding antibody responses (Figures 1 and 2A). B cells were negatively enriched from PBMCs and subjected to paired single-cell RNA and BCR sequencing (Figure 2A).

**Figure 2.**
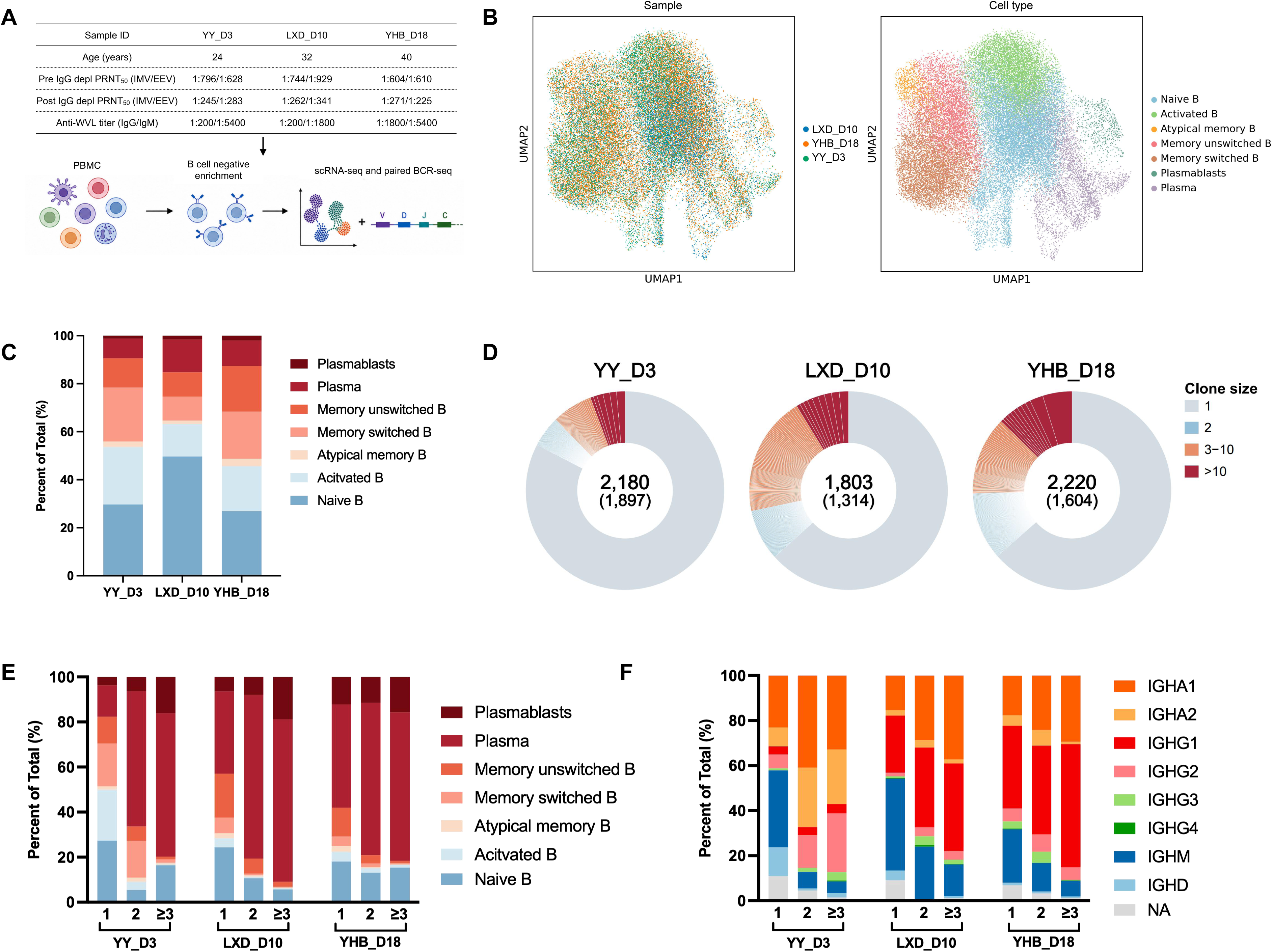
Single cell transcriptomic and BCR sequencing analysis of circulating B cells from three HIV and STI negative mpox patients. **(A)** Characteristics of the three samples selected for single B cell transcriptomic & BCR sequencing and the experimental workflow. PRNT_50_ against IMV and EEV and WVL specific IgG and IgM titers are shown as reciprocal plasma dilutions. **(B)** UMAP plots colored by sample or B cell subsets. **(C)** Proportions of major B cell subsets in each sample. **(D)** Pie charts showing BCR clonal distribution in each sample. Each sector represents an inferred clone, with colors indicating groups of different clone size; singleton clones are aggregated into a single sector. Center labels indicate total B cell numbers, with inferred clone numbers shown in parentheses. **(E)** Subset proportions of B cells stratified by clone size in each sample. **(F)** Proportions of B cells expressing different immunoglobulin isotypes among clones of different sizes. NA refers to cells with no constant-region sequence recovered by BCR sequencing.

After single-cell transcriptome quality control, 45,782 high-quality cells were retained, among which the majority were B cells (Supplementary Figures 3A and 3B). Non-B lineage cells were removed from subsequent analysis. The remaining 41,650 B cells comprised 9,944 B cells from YY_D3, 12,567 from LXD_D10 and 19,139 from YHB_D18. Unsupervised clustering and canonical marker gene expression resolved seven major B-cell states: naive B cells, activated B cells, atypical memory B cells, memory unswitched B cells, memory switched B cells, plasmablasts and plasma cells (Figure 2B and Supplementary Figure 3C). The percentage of circulating antibody secreting cells (ASCs) (plasmablasts and plasma cells) was relatively low at day 3 (YY_D3), peaked at day 10 (LXD_D10) and declined at day 18 (YHB_D18) (Figure 2C).

**Figure 3.**
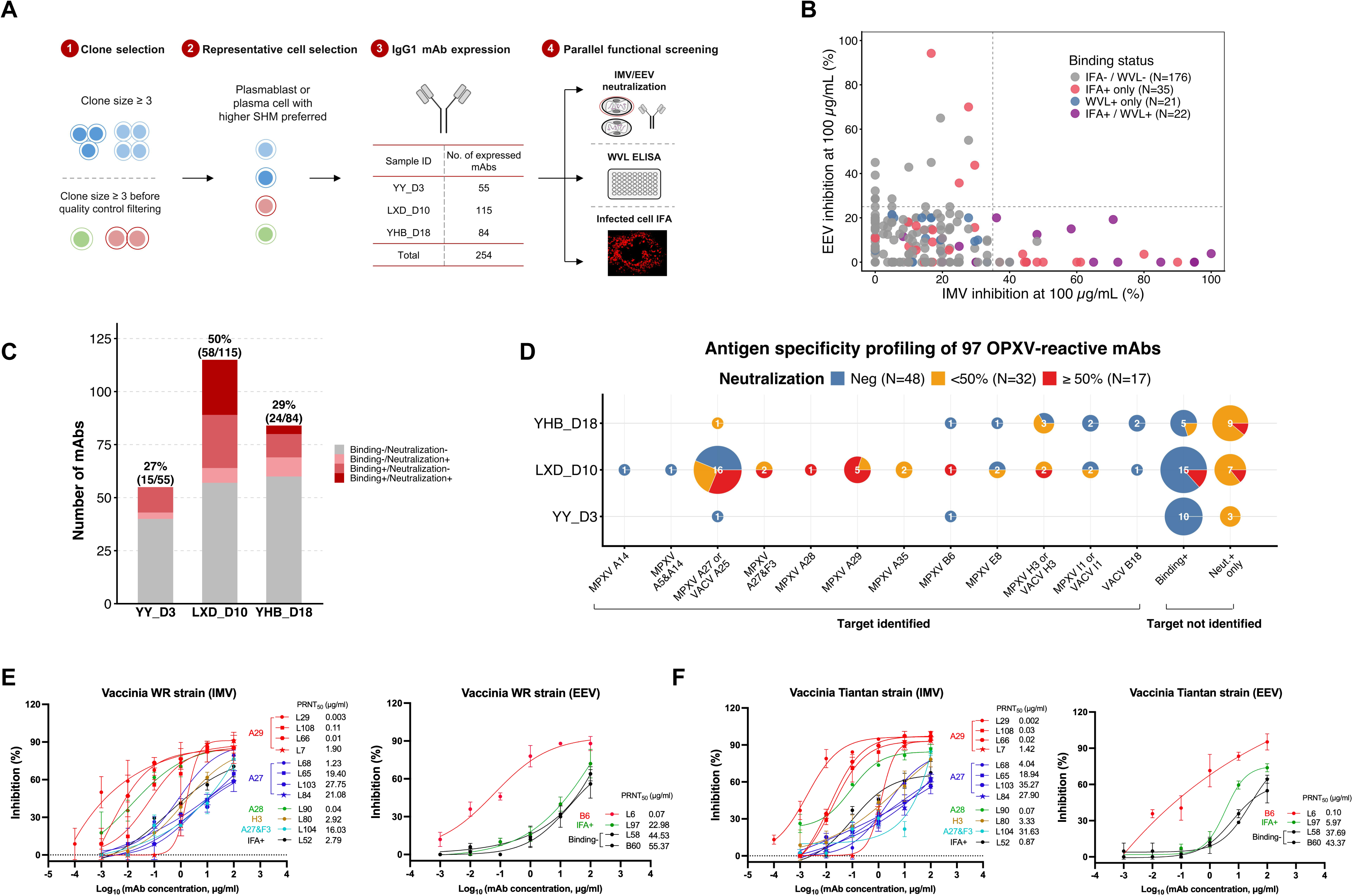
Antigen-agnostic recovery of OPXV reactive mAbs based on BCR sequencing and clonal analysis. **(A)** Workflow for clone selection and initial mAb bioactivity screening. Purified mAbs were screened via WVL based ELISA, vaccinia virus-infected cell based IFA, and in vitro IMV and EEV neutralization assays. **(B)** Functional distribution of the purified mAbs manifested in the initial bioactivity screenings. Each dot represents one mAb, and colors indicate WVL and/or IFA binding status. Dashed lines denote the thresholds defining IMV or EEV neutralization positivity. **(C)** Numbers and percentages of vaccinia virus reactive mAbs isolated from each patient, stratified by their bioactivities. Binding positivity was defined as being positive in either the WVL based ELISA or vaccinia virus-infected cell based IFA, and neutralization positivity was defined as being capable of neutralizing either IMV or EEV. **(D)** Antigen specificity profiling of 97 OPXV-reactive mAbs isolated from each patient. Bubble size indicates the number of mAbs, and fill color indicates IMV or EEV neutralizing activity at 100 μg/mL. Among mAbs whose targets remained undefined, “Binding+” denotes antibodies positive in the WVL based ELISA and/or vaccinia virus infected cell IFA, whereas “Neut.+ only” denotes antibodies positive for IMV or EEV neutralization but negative in both binding assays. **(E, F)** Titration of the neutralizing activity against vaccinia virus WR (**E**) and Tiantan (**F**) strains for purified mAbs showing an inhibition rate of ≥ 50% in the initial screening. Labels indicate identified antigen targets. For mAbs with unidentified targets, “ IFA+ ” indicates positivity in the infected cell IFA, whereas “ Binding- ” indicates negativity in both the WVL ELISA and infected cell IFA.

Acute infection may specifically stimulate germinal center responses and generate clone expanded new ASCs, meanwhile, it may also mobilize nonspecific ASCs residing in bone marrow via inflammatory signals [32, 33]. To clarify whether primary MPXV infection can drive the expansion of circulating ASCs at early time points, we next reconstructed paired immunoglobulin repertoires from productive B cells with one heavy-chain and one paired light-chain sequence. Clonotypes were assigned using Change-O based on concordant paired heavy and light chain V/J gene usage and a length normalized heavy chain CDRH3 nucleotide distance threshold of 0.185. This yielded 2,180 B cells assigned to 1,897 clones in YY_D3, 1,803 B cells assigned to 1,314 clones in LXD_D10, and 2,220 B cells assigned to 1,604 clones in YHB_D18 (Figure 2D). We compared the results of B cell clonal analysis with a virtual control sample generated by pooling together the single-cell sequencing data of PBMCs collected from 4 age and sex matched healthy donors reported in a recently study [34]. Our data show that 95.2% of B cells are singletons and only 4.8% belong to expanded clones in this virtual control sample (Supplementary Figure 4). In comparison, clone expanded cells accounted for 17.3% of BCR-recovered cells in YY_D3, 36.6% in LXD_D10 and 36.4% in YHB_D18 (Figure 2D). LXD_D10 carried the largest fraction of medium-size clones (3-10 cells per clone, 19.5%), whereas YHB_D18 contained the highest fraction of large clones (>10 cells per clone, 12.7%) (Figure 2D).

Compared with singletons, expanded clones contained higher proportions of ASCs and were enriched for class-switched isotypes (Figures 2E and 2F). IGHC usage analysis across B-cell subsets further showed that ASCs were predominantly class-switched in all three MPXV samples (Supplementary Figure 3D), with relatively higher IGHG1 usage in LXD_D10 and YHB_D18 and higher IGHA (especially IgA2) and IGHG2 usage in YY_D3 (Supplementary Figure 3D). These findings imply that acute MPXV infection may drive dynamic peripheral B cell responses.

### Antigen-agnostic antibody recovery identified mAbs targeting diverse OPXV antigens from expanded B-cell clones

The antigenic complexity of OPXV presents a methodological challenge for comprehensive mAb isolation, as antigen-defined B cell enrichment is hard to be applied to identify specific antibodies against multiple antigens simultaneously and cannot be employed to capture antibodies recognizing unknown targets. In this study, we tried to get a panoramic view of the antibody responses induced by primary acute MPXV infection via an antigen-agnostic antibody recovery strategy (Figure 3A). The rationale of this method is that antigen-reactive B cells undergo clonal selection and expansion upon infection or vaccination [35, 36], and that clonally expanded circulating ASCs are frequently enriched for antigen specific B cells [37, 38]. Clonotypes represented by at least three cells were prioritized for recombinant antibody expression (Figure 3A). For each selected clonotype, one paired heavy- and light-chain V(D)J sequence, preferably derived from an ASC with relatively high SHM, was expressed by fusing the heavy chain VDJ to the constant region of human IgG1 heavy chain and fusing the light chain VJ to a constant region of light chain selected according to its own light chain usage (Figure 3A). We also included clonotypes represented by only one or two cells if their clone sizes were ≥3 before transcriptomic and BCR quality control.

Using this strategy, we expressed 254 mAbs, including 55 from YY_D3, 115 from LXD_D10 and 84 from YHB_D18 (Figure 3A and Supplementary Table 3). The binding and neutralizing activities of the purified mAbs were screened in parallel by WVL based ELISA, vaccinia virus-infected cell-based IFA, and in vitro IMV and EEV neutralization assays (Figure 3B, Supplementary Figure 5 and Supplementary Table 3). The WVL ELISA and infected cell IFA assays are complementary to each other. Neither assay can identify all potential binding antibodies solely. We identified a total of 97 OPXV reactive mAbs, including 15 of 55 antibodies from YY_D3 (27%), 58 of 115 from LXD_D10 (50%), and 24 of 84 from YHB_D18 (29%) (Figure 3C). Among these antibodies, 48 are binding positive only, 30 are both binding and neutralizing positive and 19 are neutralizing positive only.

Next, we adopted multiple methods to determine the antigen specificity of these reactive antibodies. Initial recombinant protein-based ELISA screening identified 37 antibodies respectively reactive with MPXV A27, A29, A35, B6, E8, H3, F3, I1, A5 and A14 and VACV B18 (Supplementary Table 3). The remaining antibodies were further screened via western blotting, immunoprecipitation, and IP-MS sequentially. These analyses show that L7 recognizes MPXV A29 (Supplementary Figure 6A); L10, L30 and L26 react to VACV A25 (Supplementary Figure 6B); L80, L34, B49, and B84 bind VACV H3 (Supplementary Figures 6C and 6D); and L93 and L32 are specific for VACV I1 (Supplementary Figure 6E). Despite the above efforts, the targets of 50 reactive mAbs are still elusive at this stage, including a potent IMV neutralizing antibody (L90). Structural modeling predicted that MPXV A28 is its binding target, which was subsequently confirmed by IFA in cells transfected with a plasmid encoding MPXV A28 (Supplementary Figures 7A and 7B). L90 did not discern A28 in a denaturing western blotting assay, suggesting it recognizes a conformation sensitive epitope (Supplementary Figure 7C). Three antibodies showed dual reactivity in recombinant protein-based ELISA assays (Supplementary Table 3 and Supplementary Figure 8A). L8 recognizes both MPXV A14 and A5, whereas L81 and L104 recognize both MPXV A27 and F3. The ELISA results were confirmed by western blotting assays (Supplementary Figure 8B). Sequence alignments show limited aa sequence similarities between A14 and A5 or A27 and F3 (Supplementary Figures 8C and 8D), suggesting that the dual reactivities are unlikely due to sequence homology. Collectively, we uncovered the antigen specificities for 48 mAbs spanning 12 OPXV protein targets: MPXV A5, A14, F3, A28, A29, A35, B6, and E8; VACV B18; and the orthologous MPXV/VACV targets A27/A25, H3/H3, and I1/I1 (Figure 3D). The targets of 49 isolated OPXV-reactive antibodies remain unknown, including 27 binding-only antibodies, 19 neutralizing-only antibodies and 3 antibodies positive in both binding and neutralization assays (Figure 3D).

Moreover, antibodies that inhibited IMV or EEV infection by more than 50% at 100 μg/mL in the initial screening were further titrated through in vitro neutralization assays to determine their PRNT_50_ against vaccinia virus Tiantan and WR strains (Figures 3E and 3F). Four antibodies displayed extraordinary IMV neutralizing activities, including three A29 reactive antibodies L29, L66 and L108 and an A28 reactive antibody L90. A B6 reactive antibody, L6, demonstrated the most pronounced EEV neutralizing activity.

### The EEV neutralizing antibody conferred much stronger protection against respiratory and systemic vaccinia virus challenges than IMV neutralizing antibodies

To test whether in vitro neutralizing potency can translate into protective efficacy in vivo, three antibodies showing the most potent in vitro neutralizing activities were selected for in vivo protection experiments, including the EEV neutralizing antibody L6 and two IMV neutralizing antibodies, L29 and L66. BALB/c mice were challenged intranasally with either the WR strain (1.5×10^4^ PFU) or the Tiantan strain (6×10^5^ PFU) of vaccinia virus. Antibodies (5 mg/kg) were administered intraperitoneally 4 h before or after viral challenge (Figure 4).

**Figure 4.**
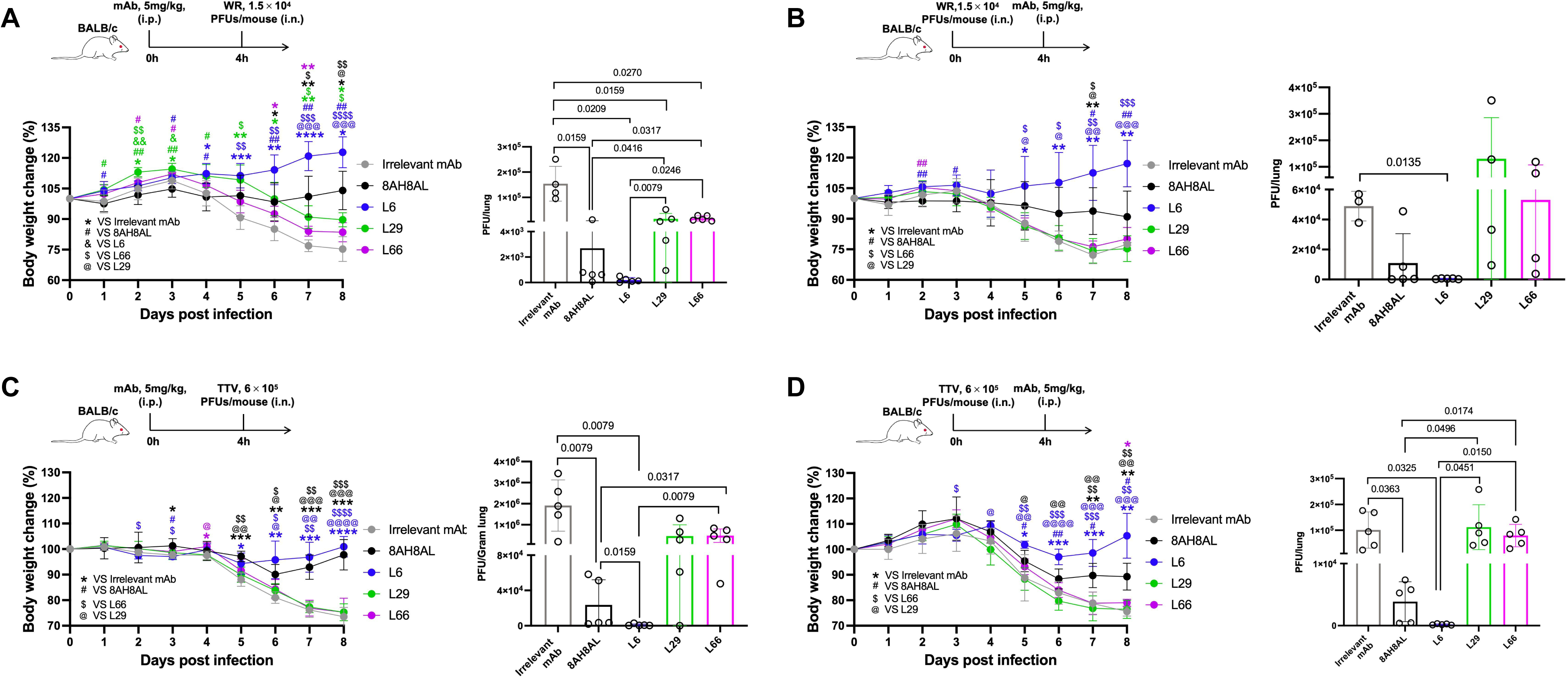
In vivo protective efficacy of mAbs showing potent in vitro neutralizing activity. BALB/c mice (n=5/group) were challenged intranasally with the WR or Tiantan strain of vaccinia virus and received a single dose (5 mg/kg) of either prophylactic or therapeutic antibody treatment. (**A, B**) Body weight changes and lung viral titers of VACV-WR challenged mice receiving prophylactic (**A**) or therapeutic (**B**) antibody treatment. (**C, D**) Body weight changes and lung viral titers of VACV-TT challenged mice receiving prophylactic (**C**) or therapeutic (**D**) antibody treatment. In the VACV-WR challenge experiment, 1 mouse died on day 8 in the group prophylactically treated with an irrelevant mAb, and 2, 1, and 1 mouse died on day 8 in groups therapeutically treated with an irrelevant mAb, L29, and L66, respectively. Lung viral titers were measured at day 8 post infection. For (**A**), (**B**), and (**D**), viral titers were determined using the whole lung and are expressed as PFU/lung, whereas for (**C**), the left lung was used and viral titers are expressed as PFU/g lung. Samples with no detectable infectious virus were assigned a value of 0. Data are shown as mean ± SD. Statistical significance is indicated as follows. *,^$^, ^#^, ^&^, ^@^: p < 0.05; **, ^$$^, ^##^, ^&&^, ^@@^: p < 0.01; ***, ^$$$^, ^###^, ^&&&^, ^@@@^: p < 0.001; ****, ^$$$$^, ^####^, ^&&&&^, ^@@@@^: p < 0.0001.

Our data show that both the prophylactic and therapeutic regimens of EEV neutralizing antibody (L6) provided nearly complete protection against weight loss caused by intranasal infection with WR strain of vaccinia (Figures 4A and 4B). Compared to an irrelevant IgG, L6 reduced lung viral titers by approximately 796-fold and 103-fold in the prophylactic and therapeutic regimens, respectively (Figures 4A and 4B). Its efficacy significantly surpassed 8AH8AL, a control antibody recognizing B6 (Figures 4A and 4B). Similar effects were also observed in mice intranasally infected with Tiantan vaccinia virus (Figures 4C and 4D). In contrast, the IMV neutralizing antibodies, L29 and L66, provided no obvious protection against intranasal Tiantan vaccinia virus infection (Figures 4C and 4D). Prophylactic administration of L29 or L66 modestly attenuated body weight loss in the WR strain intranasal challenge model and reduced lung viral titers by approximately 10.2-fold and 9.2-fold, respectively, as compared with the irrelevant antibody control (Figure 4A). While therapeutic treatment with L29 or L66 provided no obvious benefit for mice intranasally challenged with WR strain (Figure 4B). In addition, L6 also demonstrated the best protective effect against systemic TTV infection (Supplementary Figure 9). While both L6 and the IMV neutralizing antibodies (L29 and L66) tended to control viral replication, especially in testis (Supplementary Figure 9C), only L6 significantly mitigated the weight loss of mice as compared with the irrelevant control (Supplementary Figure 9A).

Fascinated by the superior in vitro and in vivo neutralizing capacities of L6, we sought to map its binding epitope. Using a phage library of random 12-mer peptides, we identified a consensus sequence that mapped to the SCR4 domain of the B6 protein (residues 220-234) (Figures 5A, 5B and 5C). The specificity of this interaction was confirmed by a peptide competition ELISA, where the B6 aa 220-234 peptide dose dependently inhibited the binding of L6 to the recombinant B6 protein but did not affect the binding by 8AH8AL (Figure 5D). These results identify a linear epitope within the SCR4 domain of B6 as the target of the highly protective EEV neutralizing antibody L6.

**Figure 5.**
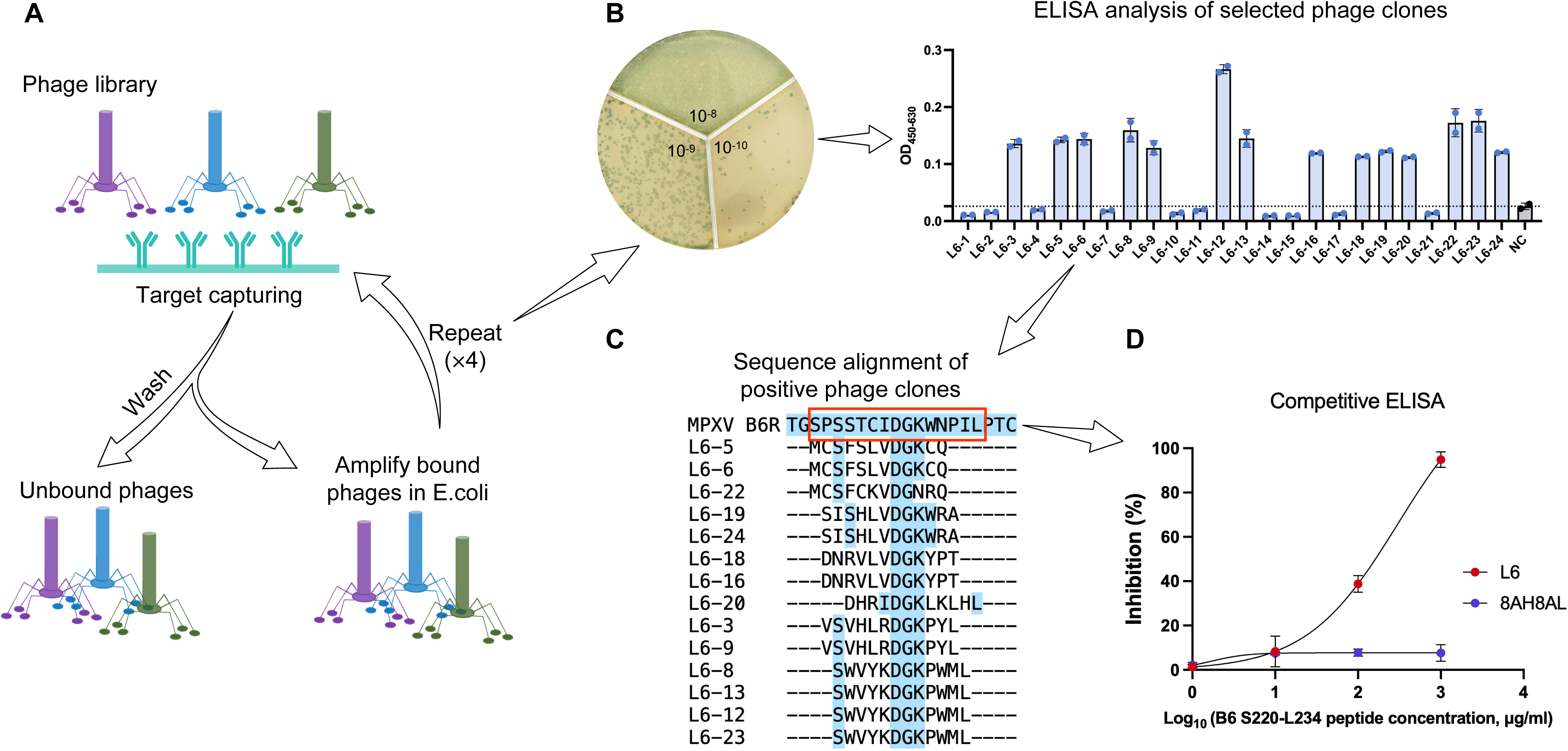
Epitope mapping for an extraordinary EEV neutralizing antibody (L6) using a phage display peptide library. **(A)** Workflow for biopanning with a random 12-mer peptide library displayed pentavalently on the M13 minor coat protein (pIII). Four rounds of binding selection, washing, elution, and amplification were performed as described in the Materials and methods. **(B)** Twenty-four phage clones were randomly selected after four rounds of biopanning and tested for binding to L6 by ELISA. Clones with OD_450-630_ values beyond the negative control (NC) were selected for Sanger sequencing. **(C)** Alignment of the peptide sequences with the MPXV B6 protein sequence. The red box highlights the peptide S220-L234 of MPXV B6 potentially recognized by mAb L6. **(D)** Competitive inhibition of L6 binding to MPXV B6 by the S220-L234 peptide. L6 and the control mAb 8AH8AL were tested at 0.5 μg/mL.

### Clonally expanded ASCs displayed distinct transcriptional states across the samples collected at different time points post symptom onset

The recovery efficiency of OPXV reactive antibodies from LXD_D10 was obviously higher than those from YY_D3 and YHB_D18, suggesting the frequency of specific circulating ASCs might change dynamically during acute MPXV infection. To understand whether the kinetics of clonally expanded circulating ASCs was linked with evolving transcriptional states, we performed pairwise differential expression analysis followed by GO enrichment analysis (Figure 6 and Supplementary Table 5) among the clonally expanded ASCs of these samples.

**Figure 6.**
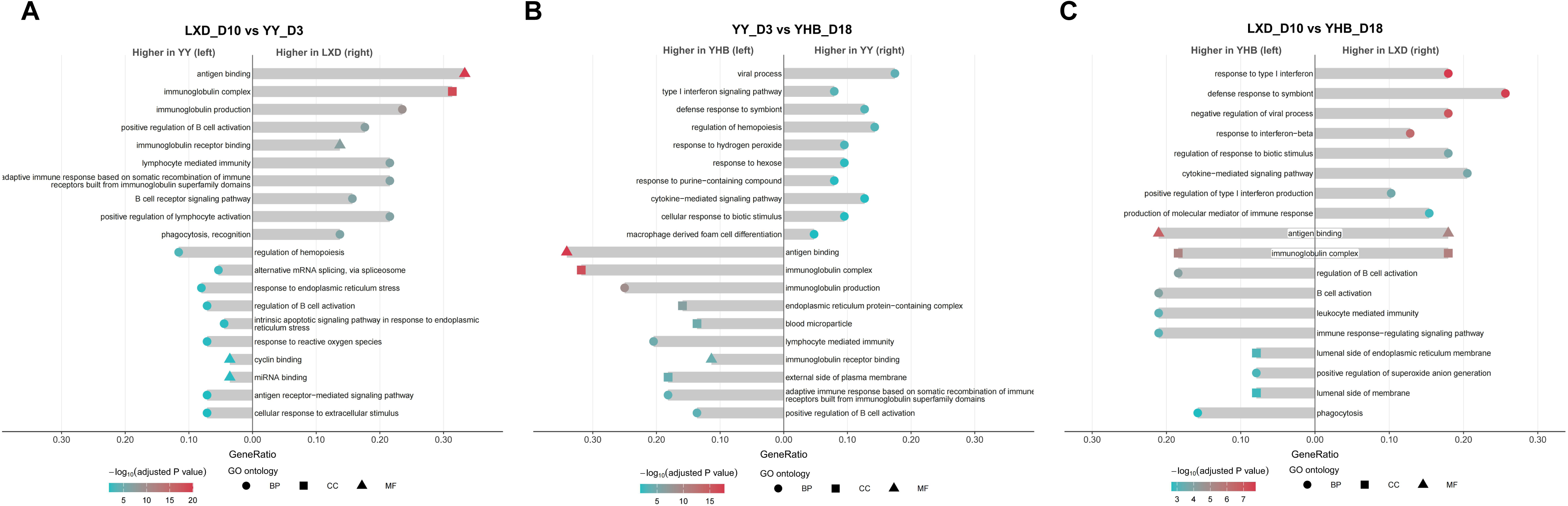
Pairwise GO enrichment analysis of clonally expanded circulating ASCs among three patients. GO enrichment analysis of differentially expressed genes identified in pairwise comparisons of clonally expanded ASCs between LXD_D10 and YY_D3 (**A**), YY_D3 and YHB_D18 (**B**), and LXD_D10 and YHB_D18 (**C**). After redundancy reduction, the top 10 representative GO terms ranked by enrichment score are shown in the figure. Bar length indicates GeneRatio, horizontal cap color represents −log_10_(adjusted P value), and cap shape denotes GO ontology: biological process (BP), cellular component (CC), or molecular function (MF).

Compared with the day 3 sample (YY_D3), the day 10 sample (LXD_D10) was enriched for antibody and B cell associated gene sets, including antigen binding, immunoglobulin complex, immunoglobulin production and B cell receptor signaling, whereas YY_D3 ASCs were enriched for hematopoietic regulation and stress or activation related pathways (Figure 6A). Compared with the day 18 sample (YHB_D18), the day 3 sample (YY_D3) upregulated antiviral and interferon related pathways, including viral process and cellular response to type I interferon, whereas YHB_D18 were enriched for antibody associated gene sets, including immunoglobulin production, immunoglobulin complex and antigen binding (Figure 6B). Differences between the day 10 (LXD_D10) and the day 18 (YHB_D18) samples are partially similar to those between day 3 and day 10 samples, with prominent interferon related and antiviral genes, including response to type I interferon, response to interferon beta and negative regulation of viral process, enriched in the earlier time point sample, whereas gene sets associated with B cell activation, leukocyte mediated immunity and immune response regulating signaling are enriched in the later time point sample (Figure 6C). In addition, our data show that the antigen binding and immunoglobulin complex related gene sets enriched in LXD_D10 and YHB_D18 ASCs are different (Supplementary Table 5). Although variations between individuals cannot be ruled out, our data indicate that the antibody secreting function increases and type I interferon signaling decreases in clonally expanded ASCs over the first 3 weeks after mpox symptom onset.

### OPXV-reactive antibody lineages recovered during acute MPXV infection showed heterogeneous SHM levels and UCA reactivity

As these patients had no known prior OPXV exposure, the recovered OPXV-reactive antibody clones offered us an opportunity to examine specific BCR clonal features during primary MPXV infection. We firstly analyzed somatic hypermutation (SHM) in clones corresponding to the 48 mAbs with verified antigen specificities (Figures 3D and 7A). These clones showed both inter- and intra-sample SHM heterogeneity. The intra-sample diversity was most apparent in LXD_D10, which yielded the largest number of antigen-defined clones (Figure 7A). In this sample, the recovered clones spanned a broad mutational range, from the unmutated clone 60 and the low-to-moderately mutated clone 18 to highly mutated lineages such as clone 30 and clone 29 (Figure 7A).

**Figure 7.**
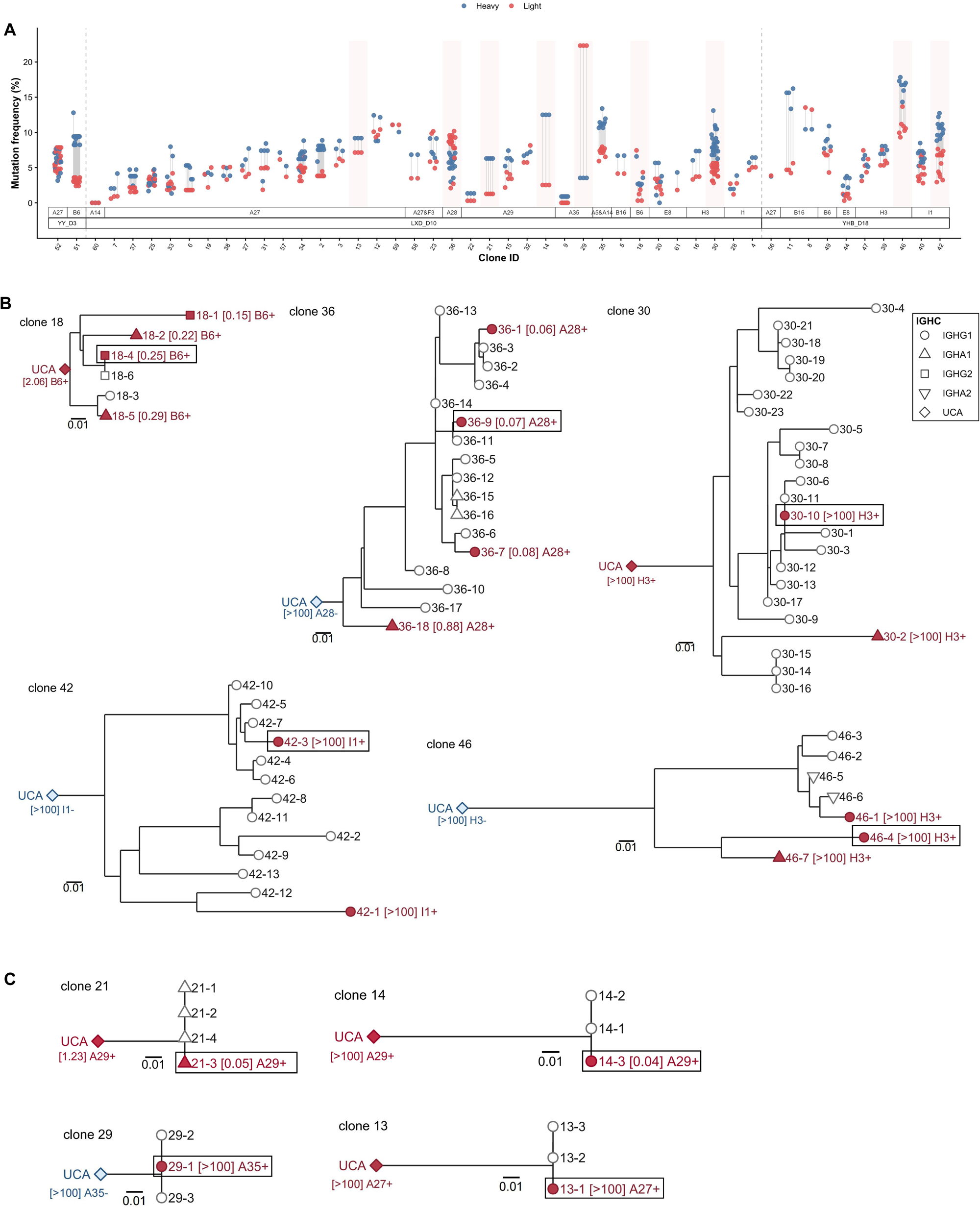
SHM and lineage analysis of OPXV-reactive antibody clones with verified antigen specificities. (**A**) SHM frequencies in the heavy- and light-chain variable regions of captured BCR sequences from clones corresponding to the 48 mAbs with verified antigen specificities. Clones are arranged by sample and antigen specificity; red shading indicates clones selected for phylogenetic analysis. (**B, C**) Heavy-chain lineage trees and functional characterization of inferred UCAs and selected lineage members from five branching lineages (**B**) and four clones whose captured members shared identical paired variable-region sequences (**C**). Inferred UCAs and selected lineage members were expressed as human IgG1 and evaluated by infected-cell based IFA, target-specific binding, and neutralizing assay. Antibodies subjected to functional evaluation are indicated by colored labels, with red and blue denoting target-specific binding-positive and binding-negative antibodies, respectively. Black boxes indicate lineage members corresponding to mAbs expressed and characterized in the initial antibody screening. Values in square brackets indicate PRNT_50_ values (μg/mL) against the corresponding IMV or EEV form of Tiantan vaccinia virus; “ >100” indicates that 50% neutralization was not reached at 100 μg/mL. Scale bars indicate genetic distance, and node shapes denote the original IGHC isotypes of the captured lineage members.

Early antibody responses during primary viral infection usually consist of low-SHM or near-germline antigen-specific antibodies [39]. In contrast, rapid expansion of highly mutated antigen-reactive B-cell lineages are commonly associated with recruitment of pre-existing memory B cells during secondary infection and vaccine recall responses [40, 41]. To further interpret the origins of antibody lineages with distinct mutational levels, we selected 9 representative clones encompassing low to relatively high SHM levels for phylogenetic analysis, including 5 branched clones (Figure 7B) and 4 clones constituted by members of identical BCR (Figure 7C). The inferred UCAs of all 9 clones and additional clonal members of the five branched lineages were expressed and evaluated for their OPXV specific binding and neutralizing activities (Supplementary Table 6 and Supplementary Figures 10-12).

Very interestingly, the UCAs showed distinct functional profiles across the 9 lineages. The UCAs of 4 clones with moderate to high somatic mutation rates displayed no measurable neutralizing or binding activity (Figures 7B and 7C), including clone 36, from which the potent A28-reactive IMV neutralizing antibody L90 was isolated. The UCAs of the other 5 clones recognized the virus, among which the UCAs of clone 18 and clone 21 exhibited pretty strong neutralizing activities, although with lower potency than their mutated descendants (Figures 7B and 7C). Notably, both clone 18 and clone 21 exhibited relatively low heavy-chain SHM levels (Figure 7A). The additional clonal members selected from the 5 branched clones showed similar functional properties to those of the initially characterized mAb from each clone and higher SHM within clones 18 and 36 was generally associated with greater neutralizing potency (Figure 7B).

### IgA1 antibodies reconstructed from L6 and L66 lineages showed limited in vitro neutralizing and in vivo protective activity

Clonal analysis showed that neutralizing antibody lineages clone 18, clone 36 and clone 21 contained clonal members of IgA1 class (Figures 7B and 7C). This was consistent with the observation that expanded clones contained a high proportion of IgA expressing B cells (Figure 2F). As early IgA antibody responses after MPXV infection are suggested to be associated with faster viral clearance from skin lesions [42], we were curious about whether the IgA antibodies found in viral specific B cell clones might provide direct protection against OPXV infection. We recovered representative IgA1 antibodies observed in clone 18 (18-2) and clone 21 (21-3) and evaluated their antiviral activities. Of note, all captured members of clone 21 were of IgA1 class and shared identical paired variable-region sequences (Figure 7C) and 21-3 was initially selected and expressed as an IgG1 antibody (L66). Both monomeric and dimeric forms of IgA1 retained their binding abilities to their cognate antigens (Supplementary Figure 13A). However, compared with their IgG1 counterparts, both IgA1 antibodies showed only weak neutralizing activity in vitro (Supplementary Figure 13B). We also evaluated the in vivo protective potential of the IgA1 antibodies using vaccinia virus infected wild type and Fcα receptor (CD89) transgene mice, respectively. Even at a high prophylactic dose of 50 mg/kg, L6-mIgA1 and L66-mIgA1 did not alleviate the weight loss of wild type or hCD89-Tg mice and failed to reduce lung viral titers (Supplementary Figures 13C and 13D). In contrast, L6 IgG1 demonstrated sterilizing protection in hCD89-Tg mice (Supplementary Figures 13C and 13D). These data suggest that specific IgA might be dispensable in MPXV containment.

## Discussion

In this study, we retrospectively investigated the natural humoral response during acute MPXV infection by integrating plasma serology, single-cell transcriptomic and BCR sequencing, circulating B cell clonal analysis and identification of OPXV reactive mAbs. All patients included in this study did not get smallpox vaccination or OPXV infection before, as China had stopped smallpox vaccination for more than 40 years before the first imported mpox case was reported in September 2022 [43]. Therefore, the results of this study faithfully represent the primary antibody responses elicited by MPXV infection.

Our data of serological tests support the view that acute MPXV infection elicited divergent IgG responses across OPXV antigens [29–31]. Although the intensities of IgG binding to 20 of 38 tested proteins correlated with IMV or EEV neutralization, individual antigen-specific responses did not fully track plasma neutralization. Meanwhile, the breadth of IgG recognition across the tested viral antigens correlated with serological neutralizing activities against both virion forms. These findings indicate that the serological antiviral activity is a combined effect of specific antibodies against multiple viral antigens. However, it is very challenging to delineate the neutralizing antibody specificity, because the neutralizing determinants of OPXV are complex [44], which can engender redundant and inter-individually heterogeneous neutralizing antibody responses [20, 45].

During the past decades, a limited number of OPXV neutralizing antibodies have been isolated from humans [13, 15, 18, 25, 46, 47], of which merely two studies tried to resolve the specificities of neutralizing antibodies in depth through either transforming PBMCs with EBV [26] or sorting circulating plasma cells for high-throughput single-cell PCR [25]. These methods were effective at isolating mAbs, but they did not allow comprehensive characterization of the cells from which the antibodies were derived. To gain deeper insight into the intra- and inter-individual diversities of antibody responses induced by acute MPXV infection, in this study, we developed a single-cell transcriptomic and BCR sequencing based antigen-agnostic mAb isolation workflow. As antigen specific B cells underwent clonal expansion can be mobilized into peripheral blood soon after infection or vaccination [48], we reason that viral specific ASCs might be enriched in the expanded B cell clones during acute MPXV infection, which may serve a good source for isolating viral specific antibodies [37, 38]. Therefore, we delineated the viral specific antibody responses adopting a strategy of recovering antibodies from each B cell clones constituted of three or more cells for three mpox patients without evidence of co-infection. Our data showed that OPXV reactive antibodies were efficiently recovered from each patient and the rank of specific antibody isolation efficiency among the three patients were in accordance with the rank of the percentages of clonally expanded B cells. The percentage of OPXV reactive clones in the sample collected at 10 days post symptom onset (LXD_D10) was astoundingly high (over 50%, 58 out of 115), while the percentages of OPXV reactive clones in samples collected at 3 days and 18 days post symptom onset were relatively low (15 out of 55 and 24 out of 84, respectively). The kinetics of MPXV specific circulating ASCs observed in this study was consistent with a previous study showing that A29-specific plasmablast frequencies peaked at day 8 post symptom onset [49] and also similar with those induced by primary vaccination and other infections [50, 51]. In addition to their frequencies, the transcriptomic status of ASCs in the sample collected at day 10 was also markedly different from those in YY_D3 and YHB_D18. Expanded ASCs from LXD_D10 were enriched for immunoglobulin-related programs relative to YY_D3 and displayed a stronger type I interferon-associated antiviral signature than YHB_D18. This transcriptional profile coincided with the highest recovery of OPXV-reactive mAbs, raising the possibility that antibody yield was influenced by the transient composition and activation state of circulating ASCs. However, because the three samples were obtained from different individuals, these observations cannot distinguish temporal effects from inter-individual variation or establish day 10 as an optimal sampling point.

Among the 97 recovered OPXV reactive mAbs, 49 showed VACV WVL binding, infected-cell IFA reactivity, and/or neutralizing activity, but their targets remained elusive. The specificities of the remaining 48 antibodies were successfully assigned using multiple methods, including recombinant viral protein-based ELISA and WB, infected or transfected cell-based IFA and IP coupled with mass spectrometry. Of note, we found that no single method was able to identify the targets for all 48 antibodies. Our findings together with two previous studies [25, 26] highlight the complexity of dissecting the specificities of antibodies elicited by natural MPXV infection, as the specificities of antibodies recognizing structure sensitive epitopes or multimeric protein complexes might be extremely hard to be defined [52]. In this study, the complex antigen-recognition pattern is exemplified by three mAbs displaying dual viral protein reactivity. As the sequence similarities are low between proteins of target pairs, we think that the observed dual reactivity is unlikely to be explained by sequence homology. Alternatively, these antibodies may recognize distinct epitopes through alternative paratope conformations, differential positioning of the epitopes within a common paratope, or engagement of overlapping paratope surfaces with distinct energetic hotspots, as demonstrated in other multispecific antibody systems [53–55].

Although the antigen specificities of 22 mAbs with measurable neutralizing activity remained unclear, we successfully defined 9 viral proteins that were recognized by neutralizing mAbs, including MPXV A27, A29, A28, A35, B6, E8, H3, I1, and F3. Neutralizing antibodies have previously been reported against the MPXV membrane proteins A29 [56], A28 [13, 15], A35 [16, 57, 58], B6 [18, 56], E8 [24], and H3 [59], as well as against the non-membrane virion core protein I1 [26]. However, to our knowledge, neutralizing antibodies recognizing MPXV A27 and F3 were isolated from mpox patients for the first time in this study. MPXV A27 was most frequently targeted, with 18 mAbs identified across all three samples, which was consistent with the serological findings of this study and previous studies [21, 60] showing that it was a prominent viral antigen recognized by mpox patients’ plasma antibodies. MPXV A27 is an A-type inclusion protein homologous to VACV A25, the latter is present in IMVs via noncovalently interacting with VACV A26 and contributes to restraining viral fusion [61]. Previous evidence for antibody-mediated inhibition targeting this protein was limited to a rabbit polyclonal anti-A25 antibody that substantially reduced VACV plaque formation in vitro [61], whereas neutralizing human mAbs against A25 or its MPXV A27 ortholog had not been reported. In this study, we found that 4 out of 18 MPXV A27-specific mAbs exhibited appreciable neutralizing activities (exceeding 50% inhibition of VACV IMV infection at 100 μg/mL), indicating that MPXV A27 specific antibodies might play a protective role during MPXV infection. Unlike MPXV A27, MPXV F3 and I1 are not present on the virion surface, but very interestingly, our data showed that one I1-reactive mAb (L93) and two A27/F3 dual-reactive mAbs (L81 and L104) could reduce the plaque formation of vaccinia virus in our in vitro screening assays. As both dual-reactive mAbs bound more strongly to F3 than A27, we reason that F3 might also be a potential neutralizing target. The exact mechanisms underlying these observations were not clarified in this study. Nonetheless, considering that MPXV F3 (a truncated homologue of the VACV E3) can block host antiviral interferon responses [62, 63] and MPXV I1 (homologue of VACV I1) is essential for the assembly of mature virions [64], we guess that antibodies targeting these proteins might inhibit virus replication via an intracellular neutralizing-like process [65].

Among the antibodies recognizing putative neutralizing determinants, three A29-reactive mAbs (L29, L66, and L108) and the A28-reactive mAb L90 showed potent IMV-neutralizing activity, whereas the B6-reactive mAb L6 showed the strongest EEV-neutralizing activity. Among these antibodies, L6, L29, and L66 were further evaluated in vivo and showed markedly different protective efficacy. L29 and L66 conferred only limited protection in the prophylactic WR respiratory challenge model, whereas L6 consistently mitigated weight loss and reduced viral burdens across prophylactic and therapeutic respiratory challenge models using the WR and Tiantan strains, as well as in the systemic Tiantan vaccinia virus infection model. Because IMVs are relatively stable and primarily mediate transmission between hosts, whereas enveloped virions promote rapid spread within the infected host, the superior protection mediated by L6 may reflect its ability to restrict EEV-mediated dissemination after the initial infection [66, 67].

Protective mAbs targeting MPXV B6 or its VACV ortholog B5 have been reported in several previous studies [18, 19, 68–71], but the epitopes have been precisely defined for only a few of them. Earlier studies positioned neutralizing epitopes to the SCR1-SCR2 region and the membrane-proximal stalk [68, 72, 73]. More recent studies suggested that mAbs recognizing the SCR3 and SCR4 regions of MPXV B6 [18, 19] could also confer protection, but the precise binding sites were not clarified. Herein, we mapped L6 to a discrete linear epitope spanning residues S220-L234 within the C-terminal portion of SCR4, immediately adjacent to the stalk region. SCR4 is required for actin-tail formation that promotes the rapid spread of enveloped virions to neighboring cells [74]. L6 binding to this region may therefore interfere with EEV dissemination efficiently, which may explain its extraordinary in vivo protective efficacy.

As aforementioned, the patients included in this study were primarily infected by MPXV, so we did not expect to isolate specific antibodies with high SHM. However, our data showed that the SHM rates were heterogeneous and a few antibody clones bore highly mutated light chain (>20%) or heavy chain (>15%). To interpret the origins of the virus specific antibody clones, we inferred the UCAs for 9 representative clones and measured their bioactivities. We found that the UCAs of 5 clones retained OPXV reactivity, while the UCAs of the other 4 clones lacked measurable binding and neutralizing activity. A recent study suggested that MPXV-specific antibodies might not be germline-encoded [15]. Our findings further propose that OPXV neutralizing antibodies may frequently originate from B cells activated by unrelated antigens prior to OPXV infection, consistent with observations in HIV infection [75]. This hypothesis is supported by our previous work showing that OPXV-reactive antibodies can be detected in individuals without a known history of smallpox vaccination or OPXV infection [76]. Collectively, these findings indicate that primary MPXV infection may either activate naïve B cells or recruit a personalized B-cell repertoire shaped by prior immune history, which may explain the highly individualized antibody specificities observed among patients encountering identical primary viral challenges.

Several neutralizing antibody clones contained members of IgA1 class, prompting us to examine whether these IgA1 antibodies retained antiviral activity. Despite the reported association between early IgA responses and faster MPXV clearance [42], IgA1 antibodies derived from the L6 and L66 lineages showed only weak in vitro neutralizing activity, and systemically administered monomeric IgA1 failed to provide protection in either wild-type or hCD89-transgenic mice.

Few limitations should be considered in this study. First, longitudinal samples were not available from the same individuals in this retrospective study, precluding longitudinal assessment of serological antibody and circulating B-cell responses for each patient. Second, the limited quantities of preserved PBMCs restricted the depth of single-cell and BCR sequencing depth, potentially leading to an underestimation of the full complexity and target diversity of the peripheral humoral response to MPXV. Third, even utilizing multiple complementary target-identification approaches, the specific targets of 49 OPXV-reactive mAbs are still elusive. Despite these limitations, our study reveals that early primary MPXV infection elicits complicated and highly dynamic humoral responses. Both naïve and nonspecifically activated B cells participate this process and contribute to the generation of OPXV neutralizing antibodies. We establish an efficient approach to deeply interrogate antibody responses induced by antigenically complex pathogens, and the neutralizing antibodies binding to unusual OPXV targets indicate further studies are warranted to fully elucidate the humoral immune responses to this viral genus.

## Materials and methods

### Ethical statement

This study was approved by the Medical Research Ethics Review Committee of Shanghai Public Health Clinical Center Affiliated to Fudan University (Approval Number: 2023-S091-02). All mouse experiments were approved by the Experimental Animal Ethics Review committee of Shanghai Public Health Clinical Center Affiliated to Fudan University (Approval number: 2025-A044-01). Experiments involving live vaccinia virus were performed under BSL-2 or ABSL-2 laboratory conditions.

### Study participants and sample collection

This retrospective study included clinical samples collected from patients diagnosed with MPXV infection and hospitalized at Shanghai Public Health Clinical Center. Residual anticoagulated peripheral blood samples obtained during routine clinical practices were reserved for this study. Demographic and clinical characteristics of the patients are summarized in Supplementary Table 1. Plasma was separated and stored at -80 °C until analysis. Peripheral blood mononuclear cells (PBMCs) were isolated by density-gradient centrifugation using Ficoll Paque Plus (Cat# 17-1440-03, GE Healthcare, USA) and preserved in liquid nitrogen until use.

### Nucleic acid extraction and plasma MPXV DNA detection by real-time PCR

Viral nucleic acids were extracted from plasma samples using the TaKaRa MiniBEST Viral RNA/DNA Extraction Kit (Cat# 9766, TaKaRa, Japan) according to the manufacturer’s instructions. MPXV DNA was detected using the Real-time PCR Detection Kit for Monkeypox Virus (Cat# SJ-XQ-018I, BioGerm, China) on an ABI 7500 real-time fluorescence quantitative PCR (qPCR) instrument (Thermo Fisher Technology Co., LTD., USA). Amplification was performed for 40 cycles. The lower detection limit of this kit was 500 copies/mL. Samples without detectable target amplification within 40 cycles were classified as not detected.

### Virus propagation and titration

IMV and EEV of vaccinia virus Tiantan and Western Reserve (ATCC VR-1354, kindly provided by Professor Xiaohui Zhou, Shanghai Public Health Clinical Center) strains were propagated and titrated as previously described [76–78]. For IMV preparation, confluent Vero cell monolayers in 100 × 10 mm culture dishes were infected with Tiantan or WR vaccinia virus at a multiplicity of infection (MOI) of 0.01. For EEV production, BHK-21 cell monolayers were infected with either virus at an MOI of 0.5. After absorption for 2 h at 37 °C in a 5% CO_2_ incubator, the inoculum was removed, and the cells were washed three times with phosphate-buffered saline (PBS). Subsequently, 8 mL of fresh maintenance medium (DMEM containing 3% FBS and 1% PS) was added to each dish, and cultures were incubated at 37 °C in a 5% CO_2_ incubator. To harvest EEV, culture supernatants were collected at 24 h post-infection and clarified by centrifugation at 800 g for 5 min at 4 °C to remove cellular debris. To harvest IMV, the infected Vero cells were lysed by three consecutive freeze-thaw cycles at 72 hours post infection.

Harvested viruses were titrated by plaque assay. Briefly, confluent monolayers of Vero cells maintained in 24-well plates were infected with 10-fold serial dilutions of IMV or EEV. An IMV-neutralizing mAb (L29, isolated and characterized in this study) was added at a final concentration of 10 μg/mL to eliminate residual IMV interference during EEV titration. After adsorption for 2 h at 37 °C in a 5% CO_2_ incubator, the inoculum was removed and cells were overlaid with 800 μL of overlay medium consisting of DMEM supplemented with 1% FBS and 1% methylcellulose. Plates were incubated at 37 °C with 5% CO_2_ for 4 days, followed by fixation with 4% paraformaldehyde and staining with 1% crystal violet. Plaques were visually counted, and viral titers were calculated and expressed as plaque-forming units (PFU) per milliliter.

### Detection of plasma binding antibodies by an in-house ELISA

Plasma binding antibodies against WVL (Tiantan strain) and OPXV proteins were measured using in-house enzyme-linked immunosorbent assay (ELISA) as previously described [76]. The recombinant protein binding assays were performed in the present study, whereas the WVL binding assay data were generated in our previous study and were reanalyzed here [27]. Briefly, high-binding 96-well ELISA plates (Cat# 9018, Corning, USA) were coated overnight at 4 °C with purified recombinant proteins at 1 μg/mL or WVL at 25 μg/mL in carbonate-bicarbonate coating buffer (30 mM NaHCO_3_, 10 mM Na_2_CO_3_, pH 9.6). The sources and catalog numbers of the recombinant OPXV proteins are listed in Supplementary Table 2.

After coating, plates were blocked with 1 × PBS containing 5% skimmed milk for 1 h at 37 °C. Plasma samples were diluted 1:100 (v/v) for the recombinant protein-based ELISA, and 3-fold serially diluted starting from 1:200 (v/v) for the WVL-based ELISA. Then, 50 μL of the diluted plasma was added to each well and incubated for 1 h at 37 °C. Plates were then washed five times with 1 × PBS containing 0.05% Tween-20. HRP-conjugated goat anti-human IgG (Cat# ab6759, Abcam, UK) or goat anti-human IgM (Cat# ab97205, Abcam, UK) secondary antibodies were added at 50 μL per well and incubated for 1 h at 37 °C. After washing, 50 μL of TMB substrate (Cat# MG882, MESGEN, China) was added and incubated for 20 min at room temperature. The reaction was stopped with 50 μL of 1 M H_2_SO_4_, and absorbances at 450nm and 630nm were measured using a microplate reader (Cat# 800TS, Biotek, USA). For each antigen, the cutoff value was defined as twice the OD_450–630_ value of a pooled negative-control plasma sample, consisting of equal volumes of plasma from six healthy individuals with no history of vaccinia vaccination.

### Plasma neutralization assay

Neutralizing activity of plasma was assessed by plaque reduction neutralization test (PRNT) using the IMV and EEV forms of vaccinia Tiantan strain. Briefly, plasma samples were 3-fold serially diluted in maintenance medium starting from 1:15 to 1:3645 (v/v).

Each diluted sample was mixed with an equal volume of virus suspension containing approximately 20 PFU per 100 µL. Virus-plasma mixtures were incubated for 1 h at 37 °C before addition to confluent Vero cell monolayers in 24-well plates. For EEV neutralization assays, the IMV-neutralizing mAb L29 was included at a final concentration of 10 µg/mL to block residual IMV infection. After incubating the virus-plasma mixtures with cells for 2 h at 37 °C in a 5% CO_2_ incubator, inocula were removed, and cells were overlaid with 800 µL of overlay medium. Plates were incubated for 4 days at 37 °C in a 5% CO_2_ incubator, fixed, and stained with crystal violet. The number of plaques in each well was visually counted. All samples were tested in duplicate, and a pooled plasma sample from six vaccinia vaccination naïve healthy individuals was included as a negative control. The viral inhibition rate was calculated as follows: Inhibition rate (%) = (1 − the average plaque number of the sample wells / the average plaque number of the negative control wells) × 100%.

### Enrichment of B cells from PBMCs

B cells were enriched from PBMCs by an immunomagnetic negative selection method using the EasySep Human Pan-B Cell Enrichment Kit (Cat# 19554, STEMCELL Technologies) according to the manufacturer’s instructions. PBMCs were incubated with antibody complexes targeting non-B-cell lineage markers, including CD2, CD3, CD14, CD16, CD36, CD42b, CD56, CD66b, CD123 and GlyA, followed by magnetic particle-mediated depletion of labeled cells. The remaining B-cell fraction was collected and resuspended in PBS for downstream single-cell analysis.

### Single-cell mRNA and V(D)J sequencing

Single-cell suspensions were resuspended in PBS at a concentration of 2 × 10^5^ cells/mL and loaded onto microwell chips using the Singleron Matrix® single-cell Processing System. Barcoding beads were collected from the microwell chips, and mRNA captured by the barcoding beads was reverse-transcribed to generate cDNA, followed by PCR amplification. A portion of the amplified cDNA was used for scRNA-seq library construction with the GEXSCOPE® single-cell RNA Library Kit, while the remaining cDNA was used for paired scBCR-seq/scTCR-seq library preparation with the sCircle single-cell Full-Length Immunoreceptor Library Kit. Immune receptor libraries were generated through cDNA circularization, digestion, purification, three rounds of TCR/BCR enrichment, fragmentation, adapter ligation, amplification, purification, and size selection. Final libraries were pooled and sequenced on an Illumina NovaSeq 6000 platform using 150-bp paired-end reads.

### Single-cell RNA-seq data processing and analysis

Raw sequencing data were processed using CeleScope v2.6.0 to generate single-cell gene-expression matrices. Cell barcodes and unique molecular identifiers (UMIs) were extracted from read 1, and read 2 was used for cDNA-based gene-expression quantification. Low-quality reads were removed, and adapter sequences and poly(A) tails were trimmed. Read 2 sequences were aligned to the human GRCh38 reference genome using STARsolo v2.7.3 [79]. PCR duplicates sharing the same cell barcode, UMI, and genomic position were removed, and UMI count matrices were generated for downstream analysis.

Raw count matrices were imported into Scanpy v1.11.3 [80] for quality control and downstream analysis. Cells were retained if they contained 500-4,000 detected genes, <15% mitochondrial genes, 5-40% ribosomal genes, and < 1% hemoglobin genes. Putative doublets were removed using Scrublet v0.2.3 [81]. Non-B-cell clusters (4,132 cells) with low CD19 and MS4A1 expression were excluded. After filtering, sample matrices were merged, yielding 41,650 high-quality B cells for downstream analysis. Counts were normalized to 10,000 UMIs per cell and log-transformed, and highly variable genes were identified. PCA was performed on highly variable genes, and sample-specific batch effects were corrected with Harmony v0.0.10 [82]. A shared nearest-neighbor graph was constructed from the corrected PCA embeddings. Cells were clustered using the Leiden algorithm at a resolution of 0.8 and visualized by UMAP.

### BCR repertoire analysis

Raw BCR sequencing data from monkeypox patients were processed using the CeleScope multi_flv_trust4 pipeline, whereas published healthy-control scBCR-seq data (samples sc_sample_29, sc_sample_30, sc_sample_31, and sc_sample_35) [34] were processed using Cell Ranger vdj v9.0.1. BCR reads were aligned to the human GRCh38 V(D)J reference for contig assembly. Productive B cells with one heavy-chain sequence and one paired light-chain sequence were retained for clonotype assignment. Productive BCR sequences were annotated using IgBLAST v1.22.0 [83] with IMGT/HighV-QUEST V(D)J gene references [84]. Initial clonotypes were assigned using Change-O v1.3.4 [85] based on shared IGHV allele usage, IGHJ allele usage, junction length, and a heavy-chain junction-distance threshold of 0.185 calculated with SHazaM v1.2.0 [86]. Heavy-chain-defined clones were further refined according to paired light-chain V and J gene usage to define final paired-chain B cell clonotypes. Somatic hypermutation frequencies were calculated using observedMutations in SHazaM, based on IgBLAST-derived germline assignments.

To infer unmutated common ancestor (UCA), paired heavy- and light-chain sequences from selected clonotypes were analyzed using Cloanalyst [87], which inferred UCA sequences and reconstructed B cell clonal lineage trees. Lineage trees were visualized in R v4.5.0 using ggplot2 v4.0.3.

### Differential expression and GO enrichment analysis of expanded ASCs

Differential gene-expression analysis and Gene Ontology (GO) enrichment analysis of clonally expanded ASCs, including plasma cells and plasmablasts, were performed using the CeleLensCloud platform at https://www.celelenscloud.cn. Genes were included in GO enrichment if they had a log_2_ fold change > 0.5, an adjusted P value < 0.05, and expression in > 10% of cells in at least one comparison group. GO enrichment results were filtered to retain terms with an adjusted P value < 0.05 and at least three genes for downstream redundancy reduction. Redundancy was reduced separately for each comparison direction and for the Biological Process, Cellular Component, and Molecular Function ontologies using rrvgo v1.20.0 in R. Pairwise semantic similarities between GO terms were calculated using the Wang method in GOSemSim v2.34.0 [88] with org.Hs.eg.db. GO terms were hierarchically clustered with a threshold of 0.7, and the term with the highest enrichment score within each cluster was retained as the representative term.

### In vitro expression and purification of mAbs

Selected paired immunoglobulin heavy- and light-chain variable region sequences were synthesized and cloned into human IgG1 or IgA1 heavy-chain and corresponding kappa or lambda light-chain expression vectors by GENEWIZ (Suzhou, China). For IgG1 expression, paired heavy and light chain plasmids were co-transfected into HEK293 cells and cultured in shake flasks at 37 °C for 5 days. Culture supernatants were harvested by centrifugation and subjected to antibody purification using an ÄKTA chromatography system with Fc affinity purification. For IgA1 expression, plasmids encoding the IgA1 heavy chain, the light chain, and the J chain were co-transfected into CHO-K1 cells and cultured in shake flasks at 37 °C for 5 days; the harvested supernatants were initially purified using a KappaXL affinity chromatography column, followed by size-exclusion chromatography (SEC) to separate and individually collect monomeric and dimeric IgA1 fractions based on molecular size. All purified antibody samples were buffer-exchanged into PBS, quantified by NanoDrop, and assessed for purity by SDS-PAGE and SEC-HPLC.

### Neutralization and binding assays of purified mAbs

PRNT assays with purified recombinant IgG1 or IgA1 mAbs were performed following the same protocol described above for plasma samples, with the exception that 3% baby rabbit complement (Cat# 31061-1, Pel-Freez Biologicals, USA) was added to the antibody-virus mixture. Isotype matched irrelevant mAbs were included as negative controls: SF-10 was used as the irrelevant control for IgG1 antibodies and monomeric or dimeric HIV-1 gp120-specific b12-IgA1 [89] was used as the irrelevant control of IgA1 antibodies. The viral inhibition rate was calculated as follows: inhibition rate (%) = (1 - the average plaque number of the antibody treated wells / the average plaque number of the negative control wells) × 100%. For antibodies screened at a single concentration of 100 μg/mL, positivity thresholds for IMV and EEV neutralization were defined as three times the inhibition rate of the purified native human IgG1 protein (Cat# ab90283, Abcam, UK) measured under the same assay conditions, corresponding to > 34.29% for IMV and > 24.00% for EEV neutralization.

Binding of mAbs to WVL or recombinant proteins was screened by ELISA following the general procedure described above for plasma samples, with all mAbs tested at a final concentration of 100 μg/mL. Recombinant OPXV proteins were firstly grouped into 9 pools and coated onto ELISA plates. Each protein pool contained 3 or 4 viral proteins and the final coating concentration for each individual protein is 0.5 μg/mL. If a positive signal was detected in a given pool, the constituent proteins were subsequently tested individually to identify the specific target antigen. The same isotype-matched irrelevant antibodies used in the neutralization assays were included as negative controls. Binding positivity was defined as an OD_450-630_ value greater than three times that of the negative control. The binding of IgA1 mAbs was detected using an HRP conjugated goat anti-human IgA alpha chain secondary antibody (Cat# ab97215, Abcam, UK).

### Immunofluorescence assay using vaccinia virus-infected cells

Vero cells were infected with vaccinia virus Tiantan strain at an MOI of 0.1 and incubated at 37 °C in a 5% CO_2_ incubator. Twenty-four hours later, infected cells were washed three times with 1 × PBS, air-dried, and fixed with pre-chilled 80% acetone at 4 °C for 10 min. Next, cells were washed with 1 × PBS and blocked with 2% bovine serum albumin (BSA) at 37 °C for 1 h. Test mAbs were added at a final concentration of 5 µg/mL and incubated for 1 h at 37 °C. Cells were washed five times with 1 × PBS and incubated with PE-conjugated anti-human IgG secondary antibody (Cat# 410708, BioLegend) for 1 h at room temperature. Fluorescence images were acquired using an inverted fluorescence microscope (Olympus CKX53, Japan). Image processing, including channel merging and brightness/contrast adjustment, was performed using ImageJ.

### MPXV A28 binding assay

Monoclonal antibody binding to MPXV A28 was assessed by immunofluorescence assay. The coding sequence of MPXV A28L (GenBank accession number: URK20576.1) fused with C-terminal 6×His tag was codon-optimized for human cell expression and cloned into a eukaryotic expression vector pJW4303 [90] by GENEWIZ (Suzhou, China).

HEK293T cells were transiently transfected with pJW4303-MPXV A28-His using Lipofectamine 2000 reagent (Cat# 11668019, Thermo Fisher Scientific, USA). Cells transfected with the empty vector were used as the negative control. At 40 h post-transfection, cells were washed three times with 1 × PBS, fixed with 4% paraformaldehyde for 15 min at room temperature, and permeabilized with 0.1% Triton X-100 for 10 min. After washing, cells were blocked with 2% BSA at 37 °C for 1 h. Cells were then co-incubated with the test mAb and mouse anti-His antibody (Cat# TA-02, ZSGB-BIO, China) for 1 h at 37 °C. After washing with 1 × PBS, cells were incubated with PE-conjugated anti-human IgG (Cat# 410708, BioLegend) and FITC-conjugated goat anti-mouse IgG (Cat# 405305, BioLegend) for 1 h at room temperature. Fluorescence images were acquired using an inverted fluorescence microscope (Olympus CKX53, Japan). Image processing, including channel merging and brightness/contrast adjustment, was performed using ImageJ.

### AlphaFold 3-based antigen-antibody complex prediction

AlphaFold 3-based antibody-antigen complex prediction was performed as previously described [13]. Full-length sequences of 40 MPXV proteins and candidate antibody heavy- and light-chain variable region sequences were used as input. For each prediction, three protein entities were defined: the MPXV protein antigen, antibody VH and antibody VL, with a stoichiometry of 1:1:1. Complex structures were predicted using the AlphaFold 3 web server [91] with default databases and presets. Predicted complex structures were visualized using PyMOL v3.0.1.

### Immunoprecipitation and western blotting

Immunoprecipitation was performed using an IP assay kit (Cat# P2180S, Beyotime, China) according to the manufacturer’s instructions. Briefly, Vero cells infected with vaccinia Tiantan virus for 24 h were washed twice with 1 × PBS and lysed in the IP lysis buffer supplemented with 10 µL protease inhibitor cocktail on ice for 1 h. Lysates were clarified by centrifugation at 10,000 g for 5 min at 4 °C, and supernatants were incubated with Protein A/G magnetic beads pre-conjugated with the indicated antibodies for 2 h at room temperature with gentle rotation. Beads were collected by magnetic separation and washed five times with the lysis buffer. Immunoprecipitated proteins were eluted by addition of 1 × SDS-PAGE loading buffer and heating at 95 °C for 5 min.

Proteins of samples were separated by SDS-PAGE using 10% or 15% PAGE gel prepared with a rapid preparation kit (Cat# PG112 or PG114, EpiZyme, Shanghai, China).

Separated proteins were then either transferred onto polyvinylidene difluoride (PVDF) membranes (Cat# IPVH00010, Millipore, USA) for immunoblotting or visualized by silver staining using the Fast Silver Stain Kit (Cat# P0017S, Beyotime, China). For western-blotting, PVDF membranes were blocked with 3% BSA in Tris-buffered saline containing 0.05% Tween-20 (TBST) for 1 h at room temperature and incubated with the test primary antibodies at 1 µg/mL. After washing with TBST, membranes were incubated with HRP-conjugated goat anti-human IgG secondary antibody (Cat# ZB-2304, ZSGB-BIO, China). Specific binding bands were developed using an ultra-sensitive ECL substrate (Cat# K-12045-D10, Advansta, USA) and captured by ChemiDoc™ Touch Imaging System (Bio-Rad, USA).

Gel bands corresponding to the molecular weights of the specific Western blot signals were excised from silver-stained gels and subjected to downstream proteomic analysis by mass spectrometry when necessary.

### Mass spectrometry analysis

Excised gel bands were reduced with 10 mM dithiothreitol for 30 min at 56 °C and alkylated with 50 mM iodoacetamide for 45 min in the dark at room temperature. Proteins were digested overnight at 37 °C with sequencing-grade modified trypsin at 5 ng/µL. Digestion was quenched with 10% formic acid, and peptides were extracted with 30% acetonitrile, dried under vacuum, and resuspended for LC-MS/MS analysis. NanoLC-MS/MS was performed using an EASY-nLC 1200 system coupled to an Orbitrap Fusion Lumos mass spectrometer (Thermo Fisher Scientific, USA). Peptides were separated on an in-house packed C18 analytical column (75 µm inner diameter × 25 cm, ReproSil-Pur 120 C18-AQ, 1.9 µm; Dr. Maisch GmbH, Germany) [92]. Peptides were eluted at a flow rate of 200 nL/min using the following gradient: 5–8% solvent B for 2 min, 8–44% solvent B for 38 min, 44–70% solvent B for 8 min, 70–100% solvent B for 2 min, and 100% solvent B for 10 min. Data were acquired in data-dependent mode, with one full MS scan followed by HCD-MS/MS scans within a 1-s cycle time. MS/MS scans were acquired in the Orbitrap using higher-energy collisional dissociation (HCD) fragmentation with an isolation window of 1.6, a resolution of 15,000, a collision energy of 30, and a maximum injection time of 22 ms. Raw MS data were searched against the UniProt protein database (Chlorocebus sabaeus and vaccinia virus) using Proteome Discoverer v2.4 with Mascot v2.7.0. The precursor and fragment mass tolerances were set to 10 ppm and 0.05 Da, respectively. Up to two missed cleavages were allowed. Carbamidomethylation of cysteine was set as a fixed modification, and protein N-terminal acetylation and methionine oxidation were set as variable modifications.

### Epitope mapping of mAb L6 by phage display

Epitope mapping of mAb L6 was performed using a Ph.D™.-12 Phage Display Peptide Library Kit v2 (Cat# E8210S, New England Biolabs, USA) according to the manufacturer’s instructions. The phage library was firstly panned using mAb L6 as the bait antibody. Briefly, L6 (10 μg/mL, 100 μL/well) was coated onto 96-well ELISA plates at 4 °C overnight. Plates were washed six times with TBST (TBS containing 0.1% Tween-20) and blocked with 0.5% bovine serum albumin (BSA, 200 μL/well) at 37 °C for 1 h. After an additional six washes with TBST, 1 × 10^11^ PFU/well (100 μL) of the Ph.D™.-12 phage library was added and incubated at room temperature for 1 h. Plates were subsequently washed ten times with TBST to remove unbound phages. Bound phages were eluted with 100 μL of elution buffer under gentle agitation for 15 min. Eluted phages were collected, titrated, and amplified according to the manufacturer’s instructions before being subjected to subsequent rounds of panning.

After four rounds of biopanning, 24 individual blue plaques were randomly picked and amplified. The amplified monoclonal phages were screened by phage ELISA using anti-M13 HRP-conjugated antibody (Cat#11973-MM05T-H, Sino Biological, China) as the secondary antibody. Wells coated with an irrelevant mAb were used as negative controls. Next, ELISA-positive clones were selected for Sanger sequencing using the sequencing primer provided in the kit (GENEWIZ, China). Peptide sequences encoded by the selected phage clones were aligned with MPXV B6 using SnapGene software.

### Peptide competition ELISA

A peptide corresponding to the sequence aa 220-234 of MPXV B6R (GenBank accession number: AAL40625.1) was synthesized (GenScript, China) at a purity of greater than 95%. Peptide competition ELISA was performed using a procedure similar to the ELISA assay described above, except that mAb L6 or the control mAb 8AH8AL [68] (0.5 µg/mL) was preincubated with serially diluted B6R aa 220-234 peptide at 37 °C for 1 h before adding to antigen-coated plates. After incubation and washing, bound antibodies were detected as described above. The percentage of competing inhibition was calculated as follows: Inhibition rate (%) = (1 - the average OD_450–630_ of peptide competition wells / the average OD_450–630_ of no-peptide control wells) × 100%.

### In vivo protection study

To evaluate protective effects of purified IgG1 mAbs against mucosal vaccinia virus infection, 4-week-old male BALB/c mice were transiently anesthetized with inhaled isoflurane (Cat# R510-22, RWD Life Science) and infected intranasally with either the vaccinia virus Tiantan strain (6 × 10^5^ PFU) or WR strain (1.5 × 10^4^ PFU) suspended in 40 µL of PBS. Monoclonal antibodies were administered intraperitoneally at 5 mg/kg in a total volume of 200 µL either 4 h before or 4 h after infection. After that, mice were monitored daily for body weight changes. On day 8 post infection, mice were euthanized and lung tissues were collected for viral titration.

To evaluate protective effects of purified IgG1 mAbs against systemic vaccinia virus infection, monoclonal antibodies were administered intraperitoneally to 4-week-old male BALB/c mice at 10 mg/kg dissolved in a total volume of 200 µL. Four hours later, the mice were challenged intravenously via the tail vein with the vaccinia virus Tiantan strain (1 × 10^7^ PFU). Body weight was monitored daily after challenge. On day 9 post infection, mice were euthanized and tissues from the lung, kidney, spleen, testis, liver, and brain were collected for viral titration.

To evaluate protective effects of monomeric IgA1 mAbs against mucosal vaccinia virus infection, 6-week-old C57BL/6 Fcα receptor (CD89) transgene mice (NM-KI-200063, Shanghai Model Organisms Center) and age-matched C57BL/6 wild-type mice were intraperitoneally treated with IgA1 mAbs at 50 mg/kg. L6-IgG1 was included as a positive control and administered intraperitoneally at 10 mg/kg. Four hours after antibody administration, mice were intranasally challenged with the vaccinia virus Tiantan strain (6 × 10^5^ PFU). Body weights were recorded daily after challenge, and mice were euthanized for lung sample collection on day 8 post infection.

### Plaque titration for vaccinia virus in mouse tissues

Vaccinia virus titers in mouse tissues were determined by a method described in our previous work [77]. Briefly, tissue samples were kept on ice and homogenized using a high-throughput tissue homogenizer (Cat# Scientz-192, NingBo Scientz Biotechnology Co., China). The homogenates were centrifuged at 2000 g for 10 minutes at 4 °C and the clarified supernatants were collected for viral titration. Briefly, confluent monolayers of Vero cells in 24-well plates were inoculated with 10-fold serially diluted tissue homogenate supernatants. After incubation, the wells were overlaid with 800 μL of overlay medium and incubated for 4 days. Finally, the plates were fixed with 4% paraformaldehyde and stained with 1% crystal violet, and plaques were visually counted.

### Statistical analysis

Statistical analyses were conducted using GraphPad Prism 10 (GraphPad Software, USA) or R version 4.5.0. AUCs were calculated in Prism using log_10_-transformed dilution factors. Ordered trends across groups were evaluated using the two-sided Jonckheere-Terpstra trend test. Correlations were assessed using two-tailed Spearman’s rank correlation analysis. Comparisons between two groups were performed using t-tests, and comparisons among multiple groups were performed using one-way ANOVA. PRNT_50_ values were calculated by four-parameter variable-slope nonlinear regression. P values < 0.05 were considered statistically significant.

## Data Availability

The data supporting the findings of this study are available from the corresponding authors upon reasonable request. The single-cell RNA-sequencing and BCR-sequencing datasets generated in this study will be deposited in a public repository upon publication of the peer-reviewed article.

## Acknowledgments

This work was supported by Shanghai Natural Science Foundation (24ZR1460500 to Z.Z.), Shanghai Oriental Top-notch Talents Program (BJWS2024088 to Z.Z.), the National Natural Science Foundation of China (32270986 to Y.W.) and Shanghai Sci-Tech Inno Center for Infection & Immunity (SSIII-202420-01 and SSIII-202420-02 to Y.W.).

## Author contributions

Y.W. and Z.Z. conceived and designed the study. Y.Z., Y.W., L.M., J.W., X.C., K.L. and R.Z. conducted most of the experiments. Y.Z., N.J., J.F. and Y.W. analyzed the data. J.W., W.Q., T.Z., R.L., H.C. and Z.Z. acquired and preserved the clinical samples. Y.Z., Y.W., J.F., and Y.R. drafted and revised the manuscript. Y.W., Z.Z. and W.Z. provided resources and funding support. All authors read and approved this version of the manuscript.

## Supplementary Figure Legends

**Supplementary Figure 1 Plasma neutralizing activities against the EEV and IMV forms of Tiantan vaccinia virus.** Plasma sample from each mpox patient was serially diluted and the neutralizing activities against IMV (**A**) and EEV (**B**) were determined by a method of PRNT. The mean inhibition rate of duplicate wells is shown for each dilution of each sample. The AUC calculated according to the neutralization curve is shown for each sample.

**Supplementary Figure 2 The time course analyses of binding IgG responses that showed weak or no significant correlations with the plasma neutralizing activities.** (**A**) Kinetics of specific IgG responses showing relatively weak correlations (r < 0.5) with the neutralizing activities. (**B**) Kinetics of specific IgG responses showing no significant correlation with the neutralizing activities. Dashed lines indicate the cutoff values defined as twice the OD_450-630_ of a negative control sample. Data are presented as medians with IQRs. Each circle represents one sample, and fill color denotes HIV status. The enlarged circles indicate the three samples selected for subsequent single-cell analysis. P values were calculated using the two-sided Jonckheere-Terpstra test for ordered trends across time windows of sample collection.

**Supplementary Figure 3 Summary of single cell sequencing data from the samples of mpox patients.** (**A**) UMAP visualization of single-cell transcriptomes without removing non-B lineage cells. (**B**) Bubble plot showing the transcription of marker genes corresponding to the cell populations shown in (**A**). (**C**) Bubble plot showing the transcription of marker genes used for B-cell subset assignment shown in Figure 2B. In (**B**) and (**C**), bubble size represents the percentage of cells expressing the gene and color intensity indicates the average transcription level. (**D**) IGHC usage across B-cell subsets in each sample. NA refers to cells with no constant-region sequence recovered by BCR sequencing.

**Supplementary Figure 4 Circulating BCR clonal analysis of healthy individuals.** The pooled pie chart was generated by combining the productive paired BCR sequences from four healthy individuals and analyzing them as a single virtual sample. Control 1 to Control 4 show the circulating BCR clonal distributions of the four healthy donors. Each sector represents an inferred clone, with colors indicating groups of different clone size; singleton clones are aggregated into a single sector. Center labels indicate total cell numbers, with inferred clone numbers shown in parentheses.

**Supplementary Figure 5 Images of IFA performed on Tiantan vaccinia virus infected cells.** Vero cells infected with vaccinia virus Tiantan strain (TTV) were incubated with indicated human mAbs, followed by detection with a PE-conjugated anti-human IgG secondary antibody. An irrelevant human mAb served as the negative control (NC).

**Supplementary Figure 6 Identification of mAb targets by IP-MS and WB.** (**A**) Identification of L7 as an MPXV A29 reactive antibody. L7 bound to MPXV A29 protein in a WB assay. The previously characterized A29 reactive mAb L29 and an irrelevant mAb were used as positive and negative controls, respectively. (**B**) Identification of L10, L30, and L26 as VACV A25 reactive antibodies. Immunoprecipitates pulled down from TTV infected or mock infected Vero cell lysates by L10, L30, L26, an anti-MPXV A27 control mAb (anti-A27 ctrl), or an irrelevant negative control mAb (NC) were analyzed by WB using the anti-MPXV A27 mAb as the primary antibody. (**C**) Identification of L80 as a VACV H3 reactive antibody. Immunoprecipitates pulled down from TTV infected or mock infected Vero cell lysates by L80 or an anti-MPXV H3 control mAb (anti-H3 ctrl) were analyzed by WB with the anti-MPXV H3 mAb as the primary antibody. (**D, E**) Identification of VACV H3 and I1 reactive antibodies. IP-MS analysis nominated VACV H3 (accession P07240) and I1 (accession P16714) as candidate targets for several antibodies (Please see Supplementary Table 4). Immunoprecipitates generated with the indicated mAbs and established H3 and I1-reactive control mAbs (anti-H3 ctrl and anti-I1 ctrl) were analyzed by WB using the anti-MPXV H3 mAb (**D**) or anti-MPXV I1 mAb (**E**) as the primary antibody. These analyses confirmed L34, B49, and B84 as H3-reactive antibodies, and L93 and L32 as I1-reactive antibodies.

**Supplementary Figure 7 Structural prediction and experimental verification of the antigen target for mAb L90.** (**A**) Predicted structural model of MPXV A28 in complex with the heavy and light chains of L90. (**B**) HEK293T cells transfected with pJW4303-MPXV A28-His or the empty vector were incubated with mAb L90 or a mouse anti-His antibody, followed by detection with a PE-conjugated anti-human IgG and an FITC-conjugated anti-mouse IgG secondary antibodies, respectively. (**C**) WB analysis of lysates from HEK293T cells transfected with pJW4303-MPXV A28-His or the empty vector. Anti-His antibody detected A28-His expression in transfected cells, whereas L90 showed no visible binding.

**Supplementary Figure 8 Verification of antigen recognition for the bispecific mAbs and the amino acid sequence alignments between the antigens recognized by the same mAb.** (**A**) Binding of the dual-reactive mAbs L81 and L104 to MPXV A27 and F3, and L8 to MPXV A5 and A14, was assessed by ELISA using recombinant proteins. (**B**) His-tagged recombinant MPXV A14, A5, A27, and F3 proteins were detected by WB using an anti-His antibody, mAbs L81, L104 and L8, or an irrelevant human mAb (NC) as the first antibody. (**C, D**) Amino acid sequence alignments of the recombinant MPXV protein fragments used for WB validation. The A14 fragment (Asn24-Ala70) was aligned with full-length A5 (Met1-Lys281) (**C**), and the A27 fragment (Leu428-Thr695) was aligned with full-length F3 (Met1-Phe153) (**D**).

**Supplementary Figure 9 Evaluation of in vivo protection against systemic Tiantan vaccinia virus infection for mAbs showing superior in vitro neutralizing capacities.** BALB/c mice (n=5/group) were treated with the indicated mAbs intraperitoneally at 10 mg/kg and challenged intravenously 4 h later with 1 x 10^7^ PFU of vaccinia virus Tiantan strain. (**A**) Experimental design and body weight change after systemic TTV infection in mice treated with L6, L29, L66, or an irrelevant IgG1 mAb. (**B, C**) Viral titers in the lung (**B**) and testis (**C**) on day 9 post-infection. Liver, spleen, kidney and brain tissues were also collected on day 9 post-infection, but infectious viral titers in these organs were all below the detection limit and are therefore not shown. One mouse in the irrelevant mAb treated group died on day 7 post-infection; therefore, viral titers were available from only 4 mice in this group. Data are shown as mean ± SD. Statistical significance is indicated as follows. *, ^@^: p < 0.05; **, ^@@^: p < 0.01.

**Supplementary Figure 10 IFA images for mAbs additionally isolated based on phylogenetic analysis.** Vero cells infected with vaccinia virus Tiantan strain were incubated with indicated human mAbs, followed by detection with a PE-conjugated anti-human IgG secondary antibody. An irrelevant human mAb was used as the negative control (NC).

**Supplementary Figure 11 Target specific binding assays for antibodies recovered from clone 30, clone 46, and clone 36.** (**A**) Immunoprecipitates pulled down from TTV infected or mock infected Vero cell lysates by MPXV H3 reactive mAb L34 were separated by SDS-PAGE and analyzed by WB using 30-2, clone 30 UCA, 46-1, 46-7, L34, or clone 46 UCA as primary antibodies. Black boxes denote specific target protein bands. (**B**) HEK293T cells transfected with pJW4303-MPXV A28-His or the empty vector were incubated with purified human mAbs or a mouse anti-His antibody, followed by detection with a PE-conjugated anti-human IgG and an FITC-conjugated anti-mouse IgG secondary antibodies, respectively. An irrelevant human mAb was included as a negative control (NC).

**Supplementary Figure 12 Determination of neutralizing titers for antibodies recovered from clone 18, clone 36, clone 21, and clone 14.** (**A**) Titration of neutralizing activities against EEV of Tiantan vaccinia virus for antibodies recovered from the MPXV B6 reactive clone 18. (**B-D**) Titration of neutralizing activities against IMV of Tiantan vaccinia virus for antibodies recovered from the MPXV A28 reactive clone 36 (**B**), A29 reactive clone 21 (**C**) and A29 reactive clone 14 (**D**).

**Supplementary Figure 13 Functional analyses of reconstructed IgA1 antibodies from L6 and L66 lineages.** (**A**) Cognate antigen binding assays for monomeric and dimeric IgA1 antibodies. The OD_450-630_ values of duplicated ELISA wells are shown for each antibody. (**B**) In vitro neutralization activities of purified IgA1 antibodies. L6-mIgA1 and L6-dIgA1 were tested against EEV of Tiantan vaccinia virus, and L66-mIgA1 and L66-dIgA1 were tested against IMV of Tiantan vaccinia virus. (**C, D**) In vivo assessment of purified IgA1 antibodies against intranasal vaccinia virus challenge in wild type and human Fcα receptor (CD89) transgene mice. Mice (n=5/group) were given L6-mIgA1 or L66-mIgA1 intraperitoneally at 50 mg/kg, or L6 IgG1 at 10 mg/kg, and challenged intranasally 4 h later with vaccinia virus Tiantan strain. An irrelevant mIgA1 was used as a negative control. Body weight changes were monitored daily (**C**), and lung viral titers were measured on day 8 post infection (**D**) and compared among groups. Data are shown as mean ± SD. One hCD89 mouse in the irrelevant mIgA1 treated group died before tissue collection on day 8; therefore, lung viral titers were available from only 4 mice in this group.

