## Supplementary Table 1 for "Decoding Humoral Immunity During Acute MPXV Infection via Comprehensive Serological Analysis and Antigen-agnostic Monoclonal Antibody Profiling"

**Supplementary Table 1 Demographic and clinical characteristics of participants**

| Characteristic | **All**  **(N = 51)** | **0–2 days (N = 6)** | **3–4 days (N = 15)** | **5–6 days (N = 7)** | **7–8 days (N = 8)** | **9–14 days (N = 7)** | **15–38 days (N = 8)** |
| --- | --- | --- | --- | --- | --- | --- | --- |
| **Age, years, median (IQR)** | 32 (28-37) | 32 (27-38) | 34 (27-37) | 32 (28-34) | 32 (31-36) | 32 (27-37) | 30 (28-39) |
| **MSM, n (%)** | 46 (90.2) | 5 (83.3) | 14 (93.3) | 7 (100.0) | 8 (100.0) | 6 (85.7) | 6 (75.0) |
| **HIV-positive, n (%)** | 20 (39.2) | 5 (83.3) | 7 (46.7) | 1 (14.3) | 3 (37.5) | 0 (0.0) | 4 (50.0) |
| **Non-HIV STI diagnosis, n (%)** | 9 (17.6) | 0 (0.0) | 4 (26.7) | 2 (28.6) | 0 (0.0) | 0 (0.0) | 3 (37.5) |
| **Onset to discharge (days), median (IQR)** | 8 (5-12) | 7 (5-12) | 10 (6-12) | 8 (7-11) | 6 (4-8) | 6 (5-10) | 18 (9-27) |
| **Plasma MPXV DNA detected, n (%)** | 24 (47.1) | 1 (16.7) | 11 (73.3) | 6 (85.7) | 0 (0.0) | 4 (57.1) | 2 (25.0) |

Note: All enrolled participants were male. Values are shown as median (IQR) or n (%). Participants were grouped according to the interval between symptom onset and sample collection. MSM, men who have sex with men; STI, sexually transmitted infection. Non-HIV STI diagnosis included syphilis and gonorrhea.
