## Supplementary Table 2 for "Decoding Humoral Immunity During Acute MPXV Infection via Comprehensive Serological Analysis and Antigen-agnostic Monoclonal Antibody Profiling"

**Supplementary Table 2 OPXV recombinant proteins used for plasma IgG binding assays**

| **Antigen category** | **Protein** | **Region expressed** | **Catalog number** | **Supplier** |
| --- | --- | --- | --- | --- |
| IMV membrane protein | MPXV A14 | Asn24-Ala70 | YVV13701 | Antibody System |
|  | MPXV A29 | Ser21-Glu110 | 40891-V08E | Sino Biological |
|  | VACV A27 | Full length | 40897-V07 | Sino Biological |
|  | VARV A30 | Full length | CSB-MP335866VAR | Cusabio |
|  | MPXV A27 | Leu428-Thr695 | YVV13001 | Antibody System |
|  | MPXV E8 | Full length | 40890-V08B | Sino Biological |
|  | VACV D8 | Met1-Thr261 | CSB-EP3211GKL1 | Cusabio |
|  | MPXV H3 | Full length | 40893-V08H1 | Sino Biological |
|  | VACV H3 | Thr21-Gly270 | CSB-EP3210GKL1 | Cusabio |
| EFC proteins | MPXV A30 | Glu28-Leu146 | YVV15001 | Antibody System |
|  | MPXV C15 | Full length | EVV17201 | Antibody System |
|  | MPXV M1 | Ala3-Gly183 | 40904-V07H | Sino Biological |
|  | VACV G9 | Gly2-Asp319 | CSB-YP357158VAI1 | Cusabio |
| EEV membrane protein | MPXV A35 | Arg58-Thr181 | 40886-V07E | Sino Biological |
|  | VACV A33 | Val57-Asn185 | 40896-V07E | Sino Biological |
|  | VARV A36 | Val57-Asn184 | CSB-MP327267VAR | Cusabio |
|  | MPXV A36 | Ile29-Lys168 | YVV12201 | Antibody System |
|  | MPXV B2 | Ser19-Asp274 | YVV12501 | Antibody System |
|  | VACV A56 | Thr17-Glu279 | CSB-EP321868VAA1 | Cusabio |
|  | MPXV B6 | Tyr18-His279 | 40902-V08H | Sino Biological |
|  | MPXV C18 | Full length | YVV12801 | Antibody System |
|  | MPXV C19 | Full length | 40894-V08B | Sino Biological |
| Viral non-membrane proteins | MPXV A33 | Thr2-Asp145 | 40885-V53E | Sino Biological |
|  | MPXV A46 | Full length | CSB-EP844975MHV | Cusabio |
|  | MPXV A5 | Full length | 40905-V07E | Sino Biological |
|  | MPXV B16 | Ile24-Glu351 | YVV17401 | Antibody System |
|  | VACV B18/B19 | Full length | 40020-V08B | Sino Biological |
|  | MPXV B5 | Asn26-Thr371 | YVV12101 | Antibody System |
|  | MPXV D13 | Full length | YVV12401 | Antibody System |
|  | MPXV D14 | Tyr20-Ala216 | YVV13601 | Antibody System |
|  | VACV C3 | Cys20-Arg263 | CSB-EP302389VAI | Cusabio |
|  | MPXV F3 | Full length | YVV14801 | Antibody System |
|  | MPXV I1 | Full length | 40888-V07E | Sino Biological |
|  | MPXV L1 | Full length | 40889-V07E | Sino Biological |
|  | MPXV A26 | Asp2-Glu75 | YVV12901 | Antibody System |
|  | MPXV A44 | Asp2-Thr74 | YVV12601 | Antibody System |
|  | MPXV B21 | Thr291-Thr487 | YVV16301 | Antibody System |
|  | VACV E6 | Ile180-Asp429 | CSB-EP325116VAI | Cusabio |

Note: Abbreviations: EEV, extracellular enveloped virion; EFC, entry-fusion complex; IMV, intracellular mature virion; MPXV, monkeypox virus; VACV, vaccinia virus; VARV, variola virus.
