## Supplementary figures for "Decoding Humoral Immunity During Acute MPXV Infection via Comprehensive Serological Analysis and Antigen-agnostic Monoclonal Antibody Profiling"

### Supplementary Figure 1

HIV negative (orange dot) · HIV positive (grey dot)

**A** IMV neutralization inhibition (%)

| Days | Sample | HIV status | 30× | 90× | 270× | 810× | 2430× | 7290× | AUC |
|---|---|---|---|---|---|---|---|---|---|
| 0–2 days | MPXV_1 D0 | positive | 67.24 | 54.95 | 20.15 | 9.05 | 20.15 | 4.95 | 66.98 |
| 0–2 days | MPXV_6 D2 | positive | 85.67 | 65.19 | 24.24 | 9.05 | 13.14 | 0.86 | 73.90 |
| 0–2 days | MPXV_17 D2 | positive | 46.76 | 32.43 | 11.10 | 24.24 | 0.00 | 0.00 | 43.49 |
| 0–2 days | MPXV_24 D2 | positive | 75.43 | 46.76 | 30.38 | 24.24 | 16.05 | 4.95 | 75.21 |
| 0–2 days | MPXV_28 D2 | negative | 16.81 | 24.37 | 9.66 | 19.75 | 4.20 | 0.00 | 31.68 |
| 0–2 days | MPXV_38 D2 | positive | 36.97 | 47.06 | 24.37 | 14.71 | 7.14 | 4.62 | 54.43 |
| 3–4 days | MPXV_5 D3 | positive | 91.81 | 71.33 | 63.14 | 32.43 | 0.00 | 0.00 | 101.50 |
| 3–4 days | MPXV_7 D3 | negative | 59.05 | 36.53 | 32.43 | 22.19 | 7.86 | 0.00 | 61.33 |
| 3–4 days | MPXV_30 D3 | negative | 74.79 | 64.71 | 39.50 | 21.85 | 2.10 | 26.89 | 85.40 |
| 3–4 days | MPXV_35 D3 | negative | 79.83 | 69.75 | 21.85 | 29.41 | 14.29 | 7.14 | 85.30 |
| 3–4 days | MPXV_40 D3 | negative | 85.67 | 32.43 | 22.19 | 28.34 | 0.00 | 4.95 | 61.20 |
| 3–4 days | MPXV_60 D3 | negative | 91.81 | 81.57 | 59.05 | 11.96 | 0.86 | 0.86 | 95.31 |
| 3–4 days | MPXV_77 D3 | positive | 73.38 | 34.48 | 15.19 | 18.10 | 0.00 | 0.00 | 49.84 |
| 3–4 days | MPXV_18 D4 | positive | 69.29 | 38.57 | 26.29 | 20.15 | 0.86 | 0.00 | 57.50 |
| 3–4 days | MPXV_19 D4 | positive | 65.19 | 44.72 | 40.62 | 20.15 | 0.00 | 0.00 | 65.88 |
| 3–4 days | MPXV_20 D4 | negative | 100.00 | 95.77 | 88.73 | 47.89 | 18.31 | 0.00 | 143.50 |
| 3–4 days | MPXV_21 D4 | positive | 73.38 | 71.33 | 26.29 | 13.14 | 11.10 | 4.95 | 76.83 |
| 3–4 days | MPXV_23 D4 | negative | 67.24 | 40.62 | 22.19 | 30.38 | 16.05 | 0.00 | 68.16 |
| 3–4 days | MPXV_26 D4 | positive | 92.44 | 64.71 | 24.37 | 19.75 | 16.81 | 6.72 | 83.60 |
| 3–4 days | MPXV_31 D4 | negative | 92.44 | 84.87 | 34.45 | 9.66 | 14.29 | 12.18 | 93.32 |
| 3–4 days | MPXV_42 D4 | positive | 85.67 | 40.62 | 5.81 | 16.05 | 13.14 | 0.00 | 56.52 |
| 5–6 days | MPXV_27 D5 | positive | 97.95 | 57.00 | 59.05 | 11.10 | 0.86 | 4.95 | 85.62 |
| 5–6 days | MPXV_11 D6 | negative | 69.75 | 57.14 | 14.29 | 7.14 | 14.71 | 0.00 | 61.15 |
| 5–6 days | MPXV_12 D6 | negative | 44.72 | 36.53 | 4.95 | 9.05 | 0.86 | 0.86 | 35.39 |
| 5–6 days | MPXV_13 D6 | negative | 92.44 | 84.87 | 67.23 | 4.62 | 16.81 | 14.71 | 108.40 |
| 5–6 days | MPXV_39 D6 | negative | 97.48 | 97.48 | 62.18 | 36.97 | 14.71 | 9.66 | 126.40 |
| 5–6 days | MPXV_44 D6 | negative | 85.67 | 73.38 | 36.53 | 26.29 | 0.00 | 2.91 | 86.11 |
| 5–6 days | MPXV_46 D6 | negative | 61.10 | 40.62 | 11.96 | 17.24 | 4.95 | 0.86 | 50.45 |
| 7–8 days | MPXV_25 D7 | positive | 89.92 | 64.71 | 21.85 | 0.00 | 16.81 | 14.71 | 74.27 |
| 7–8 days | MPXV_29 D7 | positive | 94.96 | 87.39 | 59.66 | 16.81 | 9.66 | 14.71 | 109.00 |
| 7–8 days | MPXV_36 D7 | negative | 97.48 | 100.00 | 77.31 | 54.62 | 47.06 | 24.37 | 162.20 |
| 7–8 days | MPXV_45 D7 | negative | 40.62 | 30.38 | 0.00 | 11.10 | 9.05 | 4.95 | 34.98 |
| 7–8 days | MPXV_61 D7 | negative | 100.00 | 85.67 | 87.71 | 54.95 | 11.10 | 0.00 | 138.10 |
| 7–8 days | MPXV_8 D8 | positive | 91.81 | 77.48 | 42.67 | 19.29 | 13.14 | 0.86 | 94.90 |
| 7–8 days | MPXV_32 D8 | negative | 97.48 | 92.44 | 77.31 | 59.66 | 21.85 | 19.75 | 147.80 |
| 7–8 days | MPXV_33 D8 | negative | 94.96 | 54.62 | 16.81 | 12.18 | 21.85 | 11.76 | 75.78 |
| 9–14 days | MPXV_9 D9 | negative | 71.33 | 36.53 | 28.34 | 22.19 | 2.91 | 0.00 | 59.94 |
| 9–14 days | MPXV_14 D9 | negative | 75.43 | 32.43 | 26.29 | 7.86 | 0.86 | 0.00 | 50.17 |
| 9–14 days | MPXV_43 D9 | negative | 93.86 | 42.67 | 24.24 | 17.24 | 9.05 | 0.00 | 66.86 |
| 9–14 days | MPXV_34 D10 | negative | 100.00 | 95.77 | 92.96 | 45.07 | 18.31 | 9.86 | 146.50 |
| 9–14 days | MPXV_47 D11 | negative | 93.86 | 73.38 | 36.53 | 16.05 | 22.19 | 4.95 | 94.26 |
| 9–14 days | MPXV_37 D13 | negative | 97.48 | 94.96 | 72.27 | 24.37 | 6.72 | 4.62 | 119.00 |
| 9–14 days | MPXV_10 D14 | negative | 91.81 | 71.33 | 36.53 | 11.10 | 4.95 | 0.00 | 81.02 |
| 15–38 days | MPXV_76 D16 | positive | 69.29 | 32.43 | 24.24 | 11.96 | 9.05 | 4.95 | 54.77 |
| 15–38 days | MPXV_2 D18 | negative | 100.00 | 98.59 | 84.51 | 35.21 | 4.23 | 0.00 | 130.00 |
| 15–38 days | MPXV_3 D18 | positive | 87.71 | 91.81 | 50.86 | 28.34 | 4.95 | 0.86 | 105.10 |
| 15–38 days | MPXV_64 D18 | positive | 91.81 | 69.29 | 11.96 | 20.15 | 9.05 | 4.95 | 75.77 |
| 15–38 days | MPXV_101 D20 | negative | 83.62 | 24.24 | 32.43 | 11.10 | 14.00 | 0.00 | 58.96 |
| 15–38 days | MPXV_72 D24 | positive | 100.00 | 97.95 | 75.43 | 18.10 | 0.86 | 0.00 | 115.60 |
| 15–38 days | MPXV_41 D30 | negative | 100.00 | 75.43 | 15.19 | 15.19 | 9.05 | 0.00 | 78.66 |
| 15–38 days | MPXV_69 D38 | negative | 95.90 | 91.81 | 77.48 | 50.86 | 7.86 | 2.91 | 132.40 |

Dilution

**B** EEV neutralization inhibition (%)

| Days | Sample | HIV status | 30× | 90× | 270× | 810× | 2430× | 7290× | AUC |
|---|---|---|---|---|---|---|---|---|---|
| 0–2 days | MPXV_1 D0 | positive | 76.76 | 59.86 | 42.96 | 13.38 | 5.63 | 3.52 | 77.28 |
| 0–2 days | MPXV_6 D2 | positive | 93.66 | 64.08 | 70.42 | 47.18 | 21.83 | 7.75 | 121.30 |
| 0–2 days | MPXV_17 D2 | positive | 17.61 | 7.75 | 0.00 | 17.61 | 9.86 | 14.08 | 24.36 |
| 0–2 days | MPXV_24 D2 | positive | 74.65 | 47.18 | 13.38 | 26.06 | 16.20 | 1.41 | 67.20 |
| 0–2 days | MPXV_28 D2 | negative | 15.49 | 3.52 | 0.00 | 0.00 | 0.00 | 0.00 | 5.38 |
| 0–2 days | MPXV_38 D2 | positive | 13.38 | 9.15 | 2.82 | 0.00 | 0.00 | 0.00 | 8.90 |
| 3–4 days | MPXV_5 D3 | positive | 89.44 | 87.32 | 45.07 | 36.62 | 9.15 | 26.06 | 112.60 |
| 3–4 days | MPXV_7 D3 | negative | 80.99 | 55.63 | 17.61 | 30.28 | 7.75 | 3.52 | 73.25 |
| 3–4 days | MPXV_30 D3 | negative | 89.44 | 47.18 | 0.00 | 0.00 | 0.00 | 0.00 | 43.85 |
| 3–4 days | MPXV_35 D3 | negative | 57.75 | 15.49 | 0.00 | 1.41 | 0.00 | 0.00 | 21.84 |
| 3–4 days | MPXV_40 D3 | negative | 74.65 | 78.87 | 34.51 | 0.00 | 0.00 | 0.00 | 71.90 |
| 3–4 days | MPXV_60 D3 | negative | 89.44 | 76.76 | 38.73 | 19.72 | 11.97 | 11.27 | 94.25 |
| 3–4 days | MPXV_77 D3 | positive | 85.21 | 61.97 | 17.61 | 16.20 | 3.52 | 11.97 | 70.56 |
| 3–4 days | MPXV_18 D4 | positive | 74.65 | 38.73 | 0.00 | 7.75 | 19.72 | 9.86 | 51.74 |
| 3–4 days | MPXV_19 D4 | positive | 76.76 | 51.41 | 23.94 | 3.52 | 21.83 | 1.41 | 66.70 |
| 3–4 days | MPXV_20 D4 | negative | 95.48 | 93.67 | 73.76 | 42.08 | 27.60 | 7.69 | 137.70 |
| 3–4 days | MPXV_21 D4 | positive | 76.76 | 53.52 | 19.72 | 28.17 | 0.00 | 11.27 | 69.38 |
| 3–4 days | MPXV_23 D4 | negative | 80.99 | 49.30 | 30.28 | 11.97 | 13.38 | 7.75 | 71.23 |
| 3–4 days | MPXV_26 D4 | positive | 72.54 | 47.18 | 34.51 | 1.41 | 0.00 | 7.75 | 58.80 |
| 3–4 days | MPXV_31 D4 | negative | 72.54 | 57.75 | 42.96 | 3.52 | 5.63 | 5.63 | 71.06 |
| 3–4 days | MPXV_42 D4 | positive | 91.55 | 74.65 | 28.17 | 14.08 | 7.75 | 0.00 | 81.31 |
| 5–6 days | MPXV_27 D5 | positive | 83.10 | 76.76 | 49.30 | 0.00 | 0.00 | 0.00 | 79.97 |
| 5–6 days | MPXV_11 D6 | negative | 72.54 | 15.49 | 0.00 | 0.00 | 0.00 | 0.00 | 24.69 |
| 5–6 days | MPXV_12 D6 | negative | 14.08 | 9.15 | 0.00 | 11.97 | 17.61 | 11.97 | 24.69 |
| 5–6 days | MPXV_13 D6 | negative | 78.87 | 80.99 | 17.61 | 0.00 | 0.00 | 0.00 | 65.85 |
| 5–6 days | MPXV_39 D6 | negative | 83.10 | 76.76 | 57.75 | 23.94 | 3.52 | 7.75 | 98.95 |
| 5–6 days | MPXV_44 D6 | negative | 91.55 | 66.20 | 21.83 | 18.31 | 3.52 | 13.38 | 77.45 |
| 5–6 days | MPXV_46 D6 | negative | 72.54 | 51.41 | 32.39 | 1.41 | 3.52 | 0.00 | 59.64 |
| 7–8 days | MPXV_25 D7 | positive | 80.99 | 57.75 | 5.63 | 0.00 | 0.00 | 3.52 | 50.40 |
| 7–8 days | MPXV_29 D7 | positive | 93.66 | 93.66 | 59.86 | 13.38 | 0.00 | 0.00 | 102.00 |
| 7–8 days | MPXV_36 D7 | negative | 95.77 | 91.55 | 72.54 | 11.27 | 1.41 | 1.41 | 107.50 |
| 7–8 days | MPXV_45 D7 | negative | 9.86 | 11.27 | 7.75 | 3.52 | 21.83 | 13.38 | 26.71 |
| 7–8 days | MPXV_61 D7 | negative | 87.32 | 68.31 | 70.42 | 13.38 | 9.86 | 11.97 | 101.00 |
| 7–8 days | MPXV_8 D8 | positive | 93.66 | 74.65 | 61.97 | 19.72 | 5.63 | 5.63 | 101.00 |
| 7–8 days | MPXV_32 D8 | negative | 100.00 | 100.00 | 72.54 | 11.97 | 7.75 | 0.00 | 115.60 |
| 7–8 days | MPXV_33 D8 | negative | 91.55 | 80.99 | 51.41 | 4.93 | 11.97 | 0.00 | 93.07 |
| 9–14 days | MPXV_9 D9 | negative | 76.76 | 49.30 | 1.41 | 1.41 | 9.86 | 13.38 | 51.07 |
| 9–14 days | MPXV_14 D9 | negative | 85.21 | 40.85 | 0.00 | 5.63 | 0.00 | 0.00 | 42.50 |
| 9–14 days | MPXV_43 D9 | negative | 83.10 | 80.99 | 74.65 | 49.30 | 45.07 | 11.27 | 141.80 |
| 9–14 days | MPXV_34 D10 | negative | 96.38 | 95.48 | 85.52 | 50.23 | 27.60 | 0.45 | 146.60 |
| 9–14 days | MPXV_47 D11 | negative | 74.65 | 55.63 | 57.75 | 30.28 | 17.61 | 11.27 | 97.44 |
| 9–14 days | MPXV_37 D13 | negative | 93.66 | 78.87 | 53.52 | 9.86 | 5.63 | 0.00 | 92.91 |
| 9–14 days | MPXV_10 D14 | negative | 80.99 | 72.54 | 47.18 | 18.31 | 26.06 | 3.52 | 98.45 |
| 15–38 days | MPXV_76 D16 | positive | 72.54 | 55.63 | 7.75 | 11.27 | 16.20 | 14.08 | 64.01 |
| 15–38 days | MPXV_2 D18 | negative | 100.00 | 92.76 | 81.90 | 37.56 | 16.74 | 0.00 | 133.10 |
| 15–38 days | MPXV_3 D18 | positive | 89.44 | 70.42 | 55.63 | 19.72 | 17.61 | 11.27 | 102.00 |
| 15–38 days | MPXV_64 D18 | positive | 87.32 | 78.87 | 23.94 | 17.61 | 2.82 | 0.00 | 79.63 |
| 15–38 days | MPXV_101 D20 | negative | 89.44 | 64.08 | 40.85 | 19.72 | 26.06 | 16.20 | 97.10 |
| 15–38 days | MPXV_72 D24 | positive | 80.99 | 72.54 | 53.52 | 17.61 | 23.94 | 16.20 | 103.20 |
| 15–38 days | MPXV_41 D30 | negative | 89.44 | 78.87 | 53.52 | 9.15 | 13.38 | 9.86 | 97.61 |
| 15–38 days | MPXV_69 D38 | negative | 76.76 | 80.99 | 55.63 | 21.83 | 1.41 | 11.97 | 97.44 |

Dilution

### Supplementary Figure 2

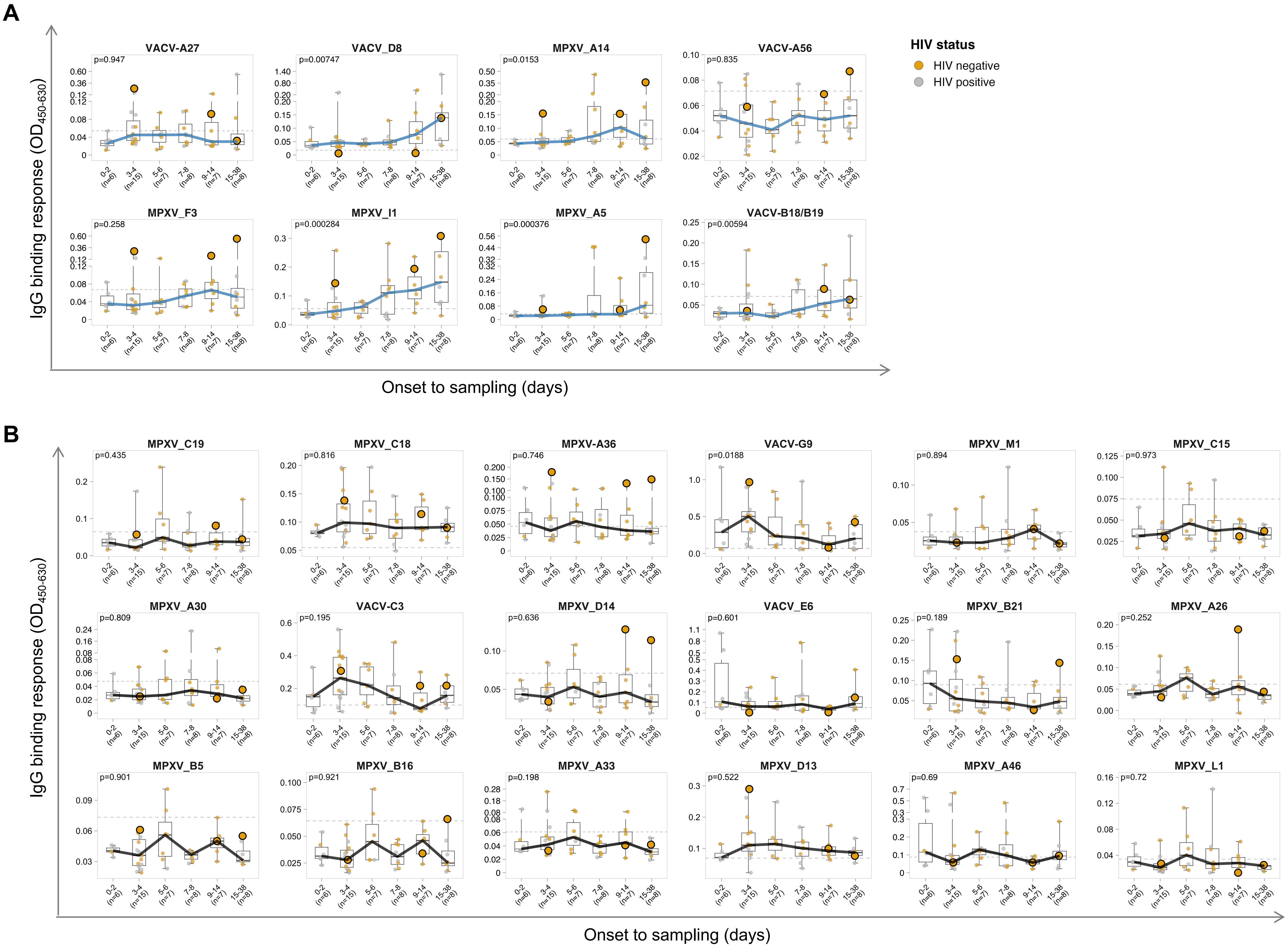

### Supplementary Figure 3

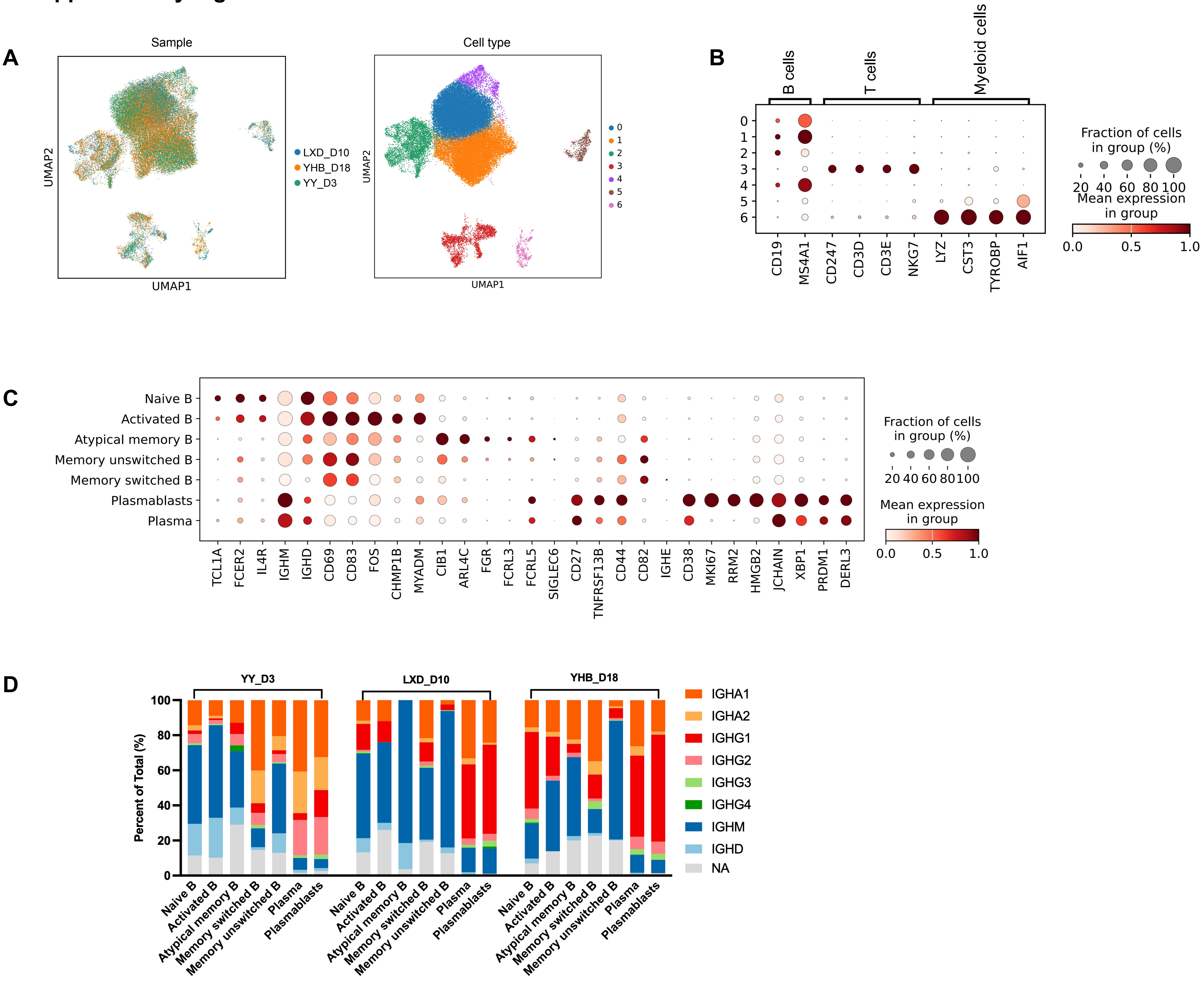

### Supplementary Figure 4

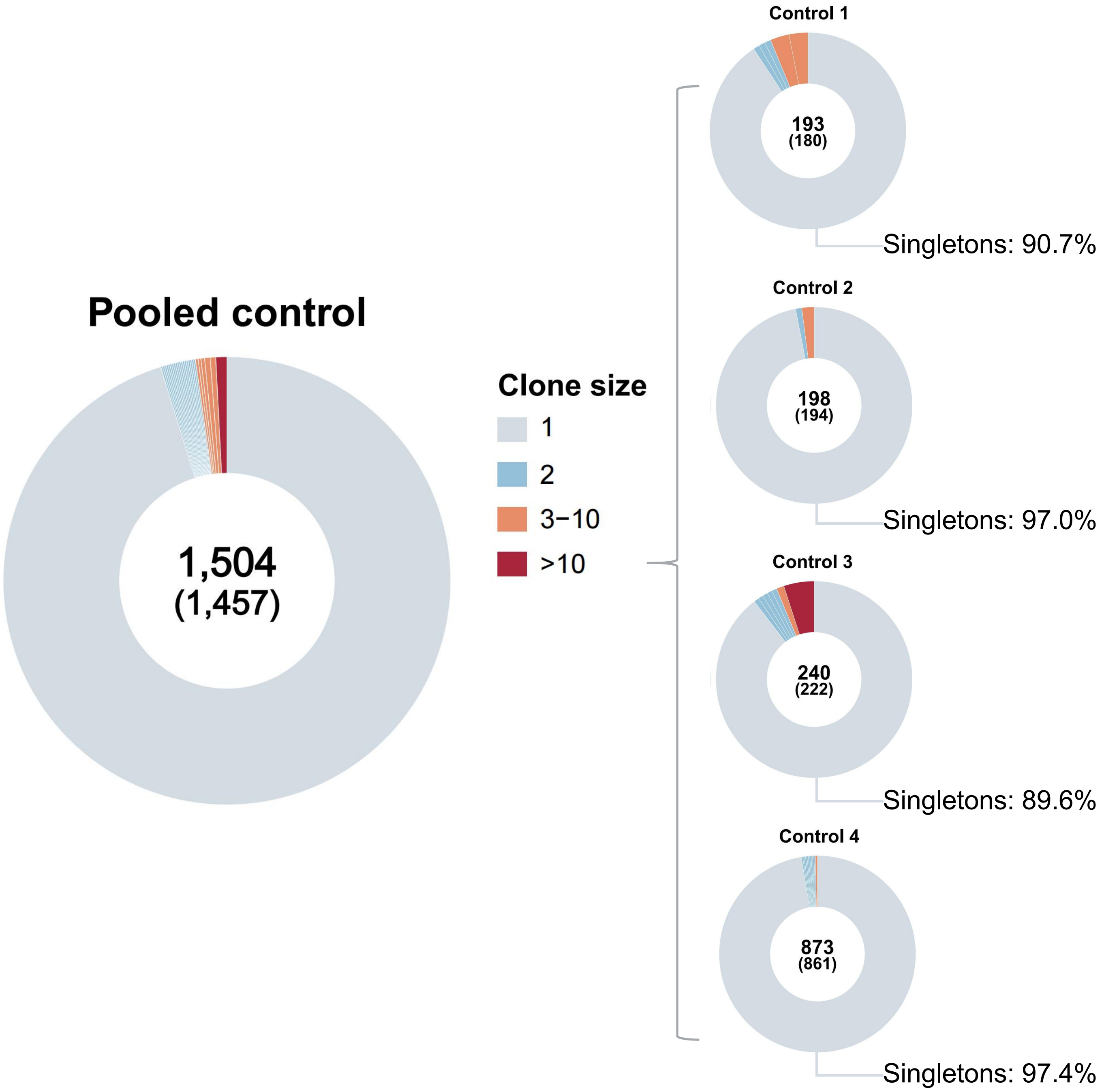

#### Supplementary Figure 5

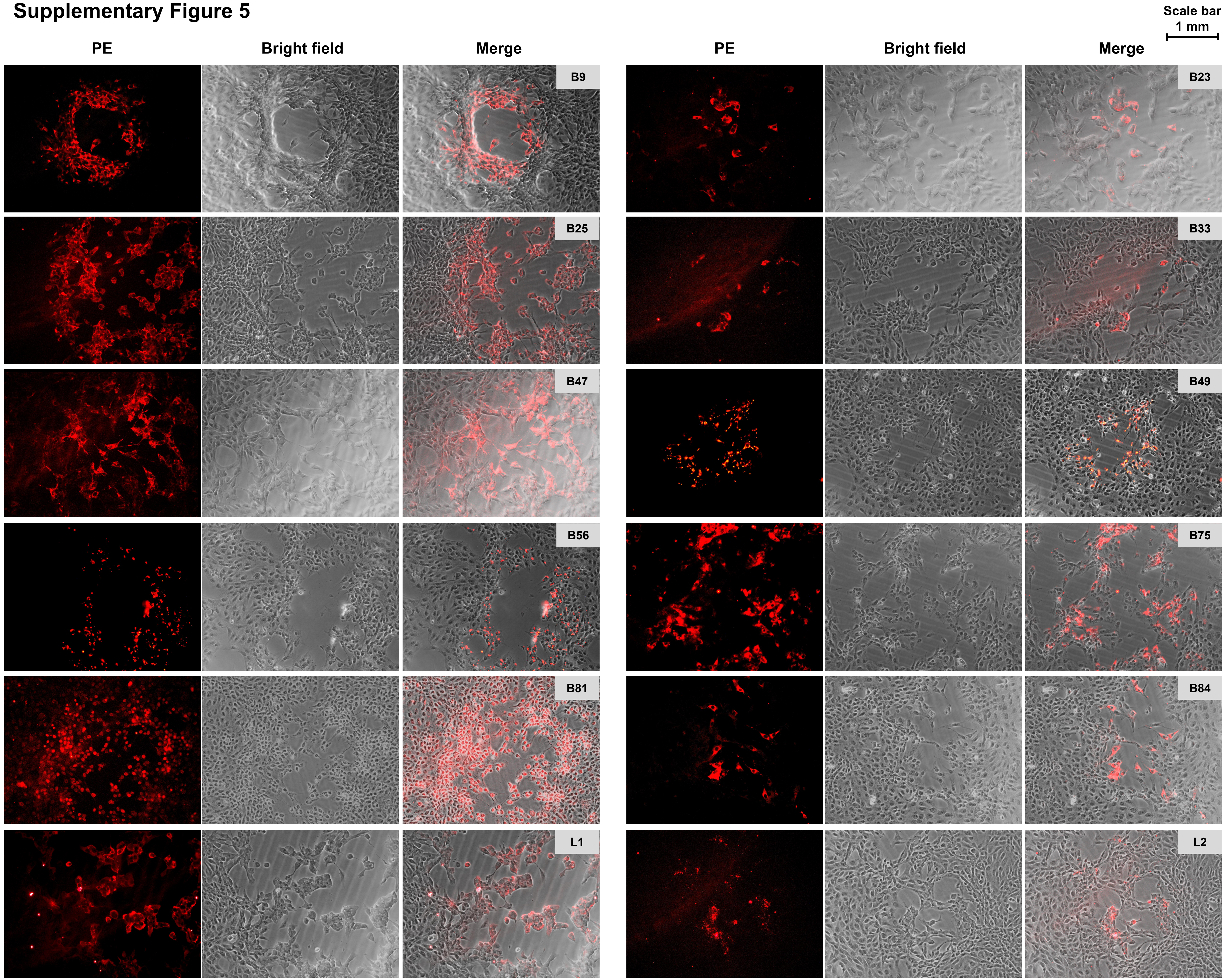

#### Supplementary Figure 5

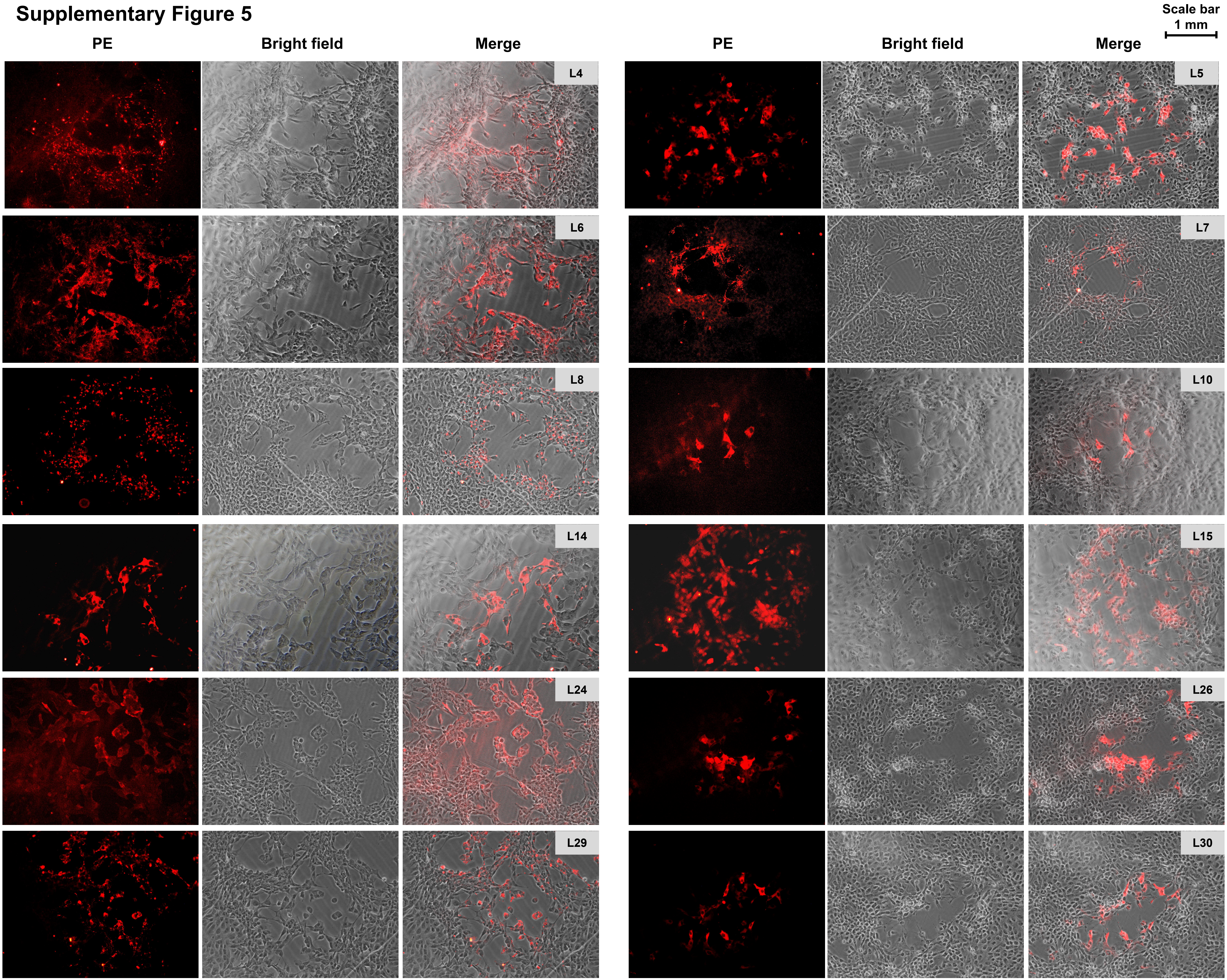

#### Supplementary Figure 5

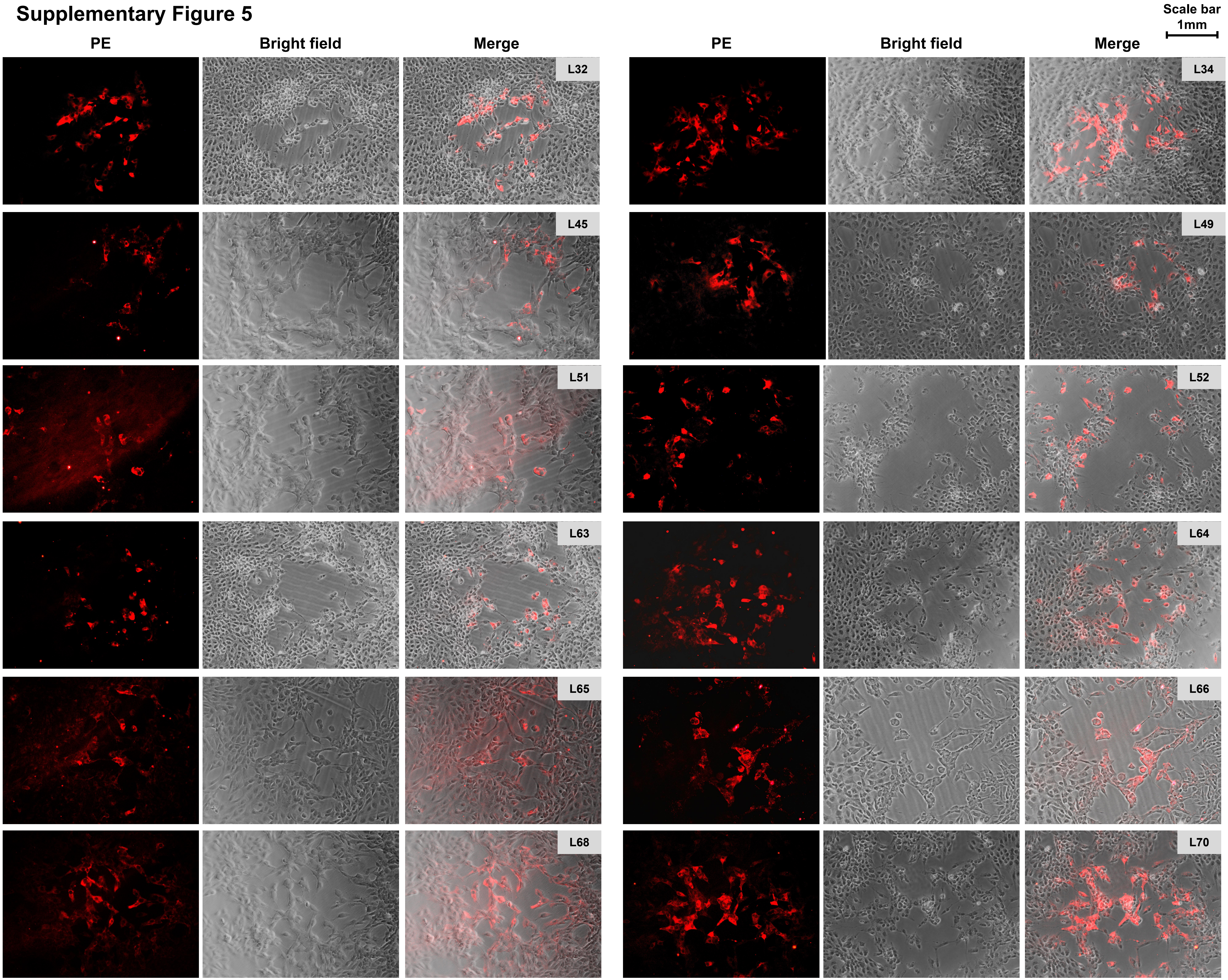

#### Supplementary Figure 5

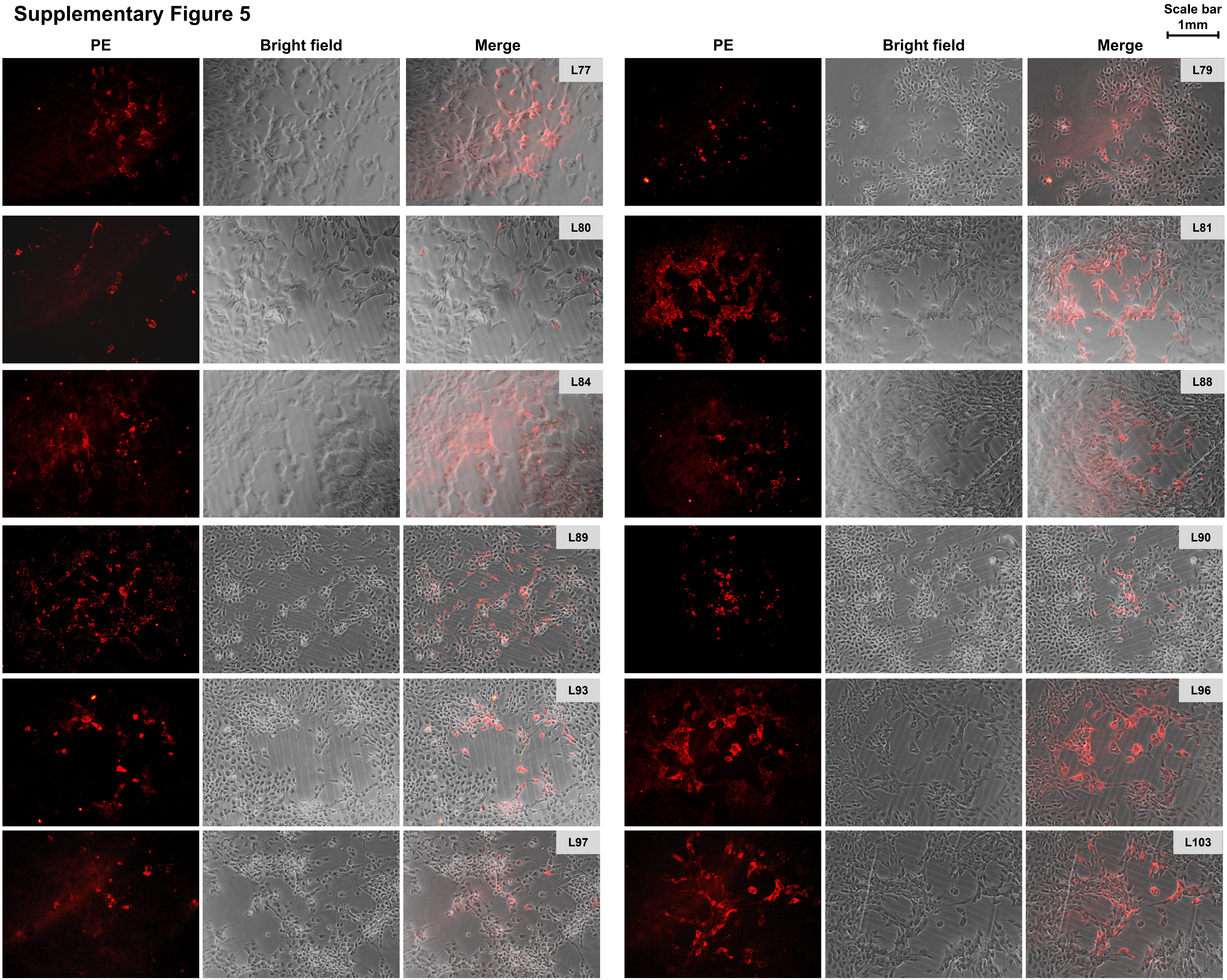

### Supplementary Figure 5

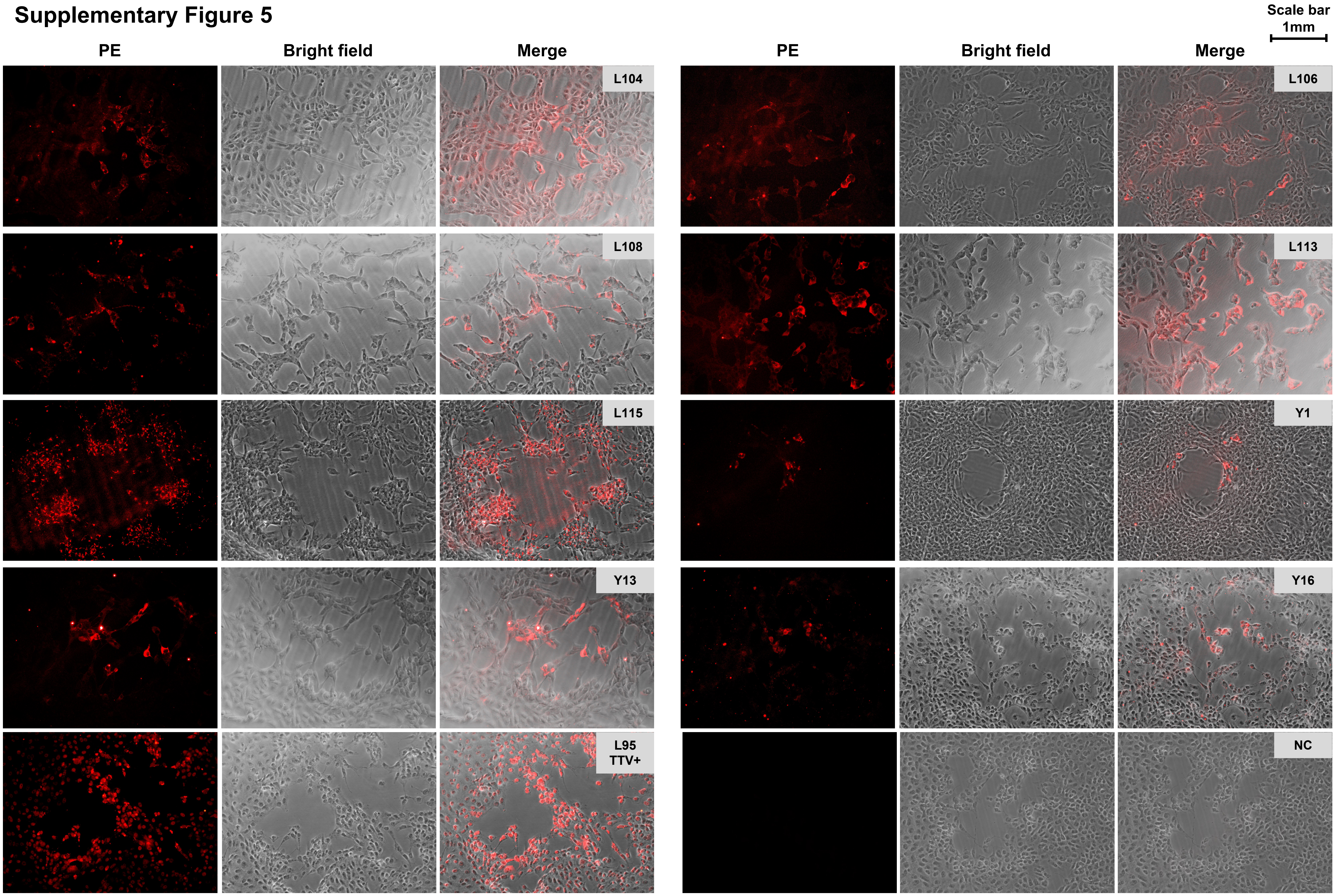

### Supplementary Figure 6

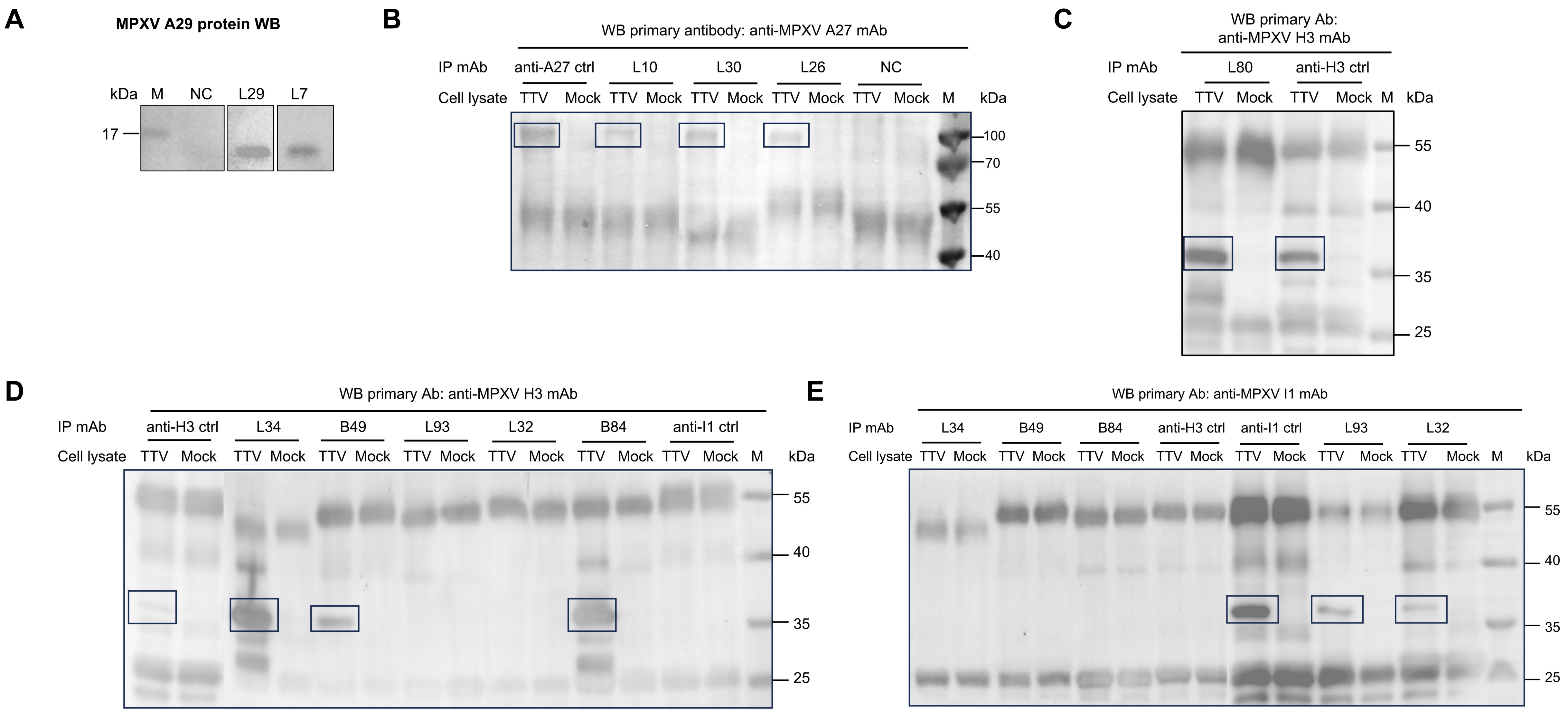

#### Supplementary Figure 7

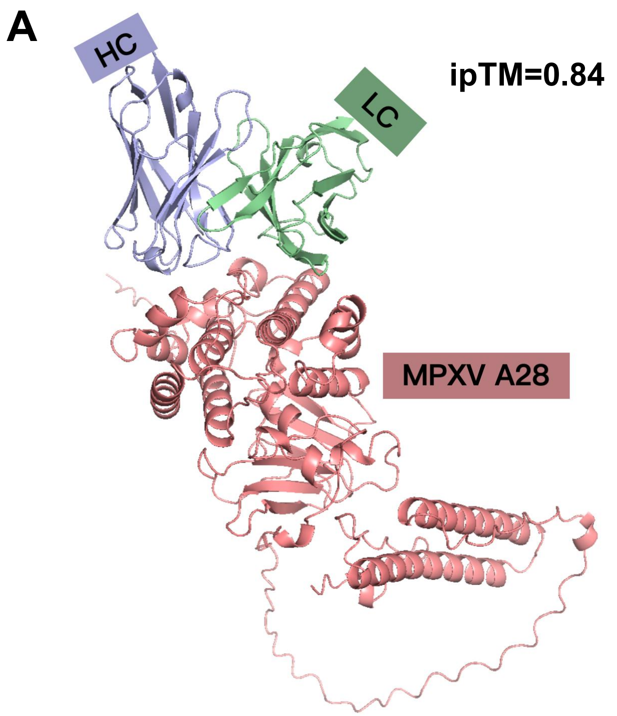


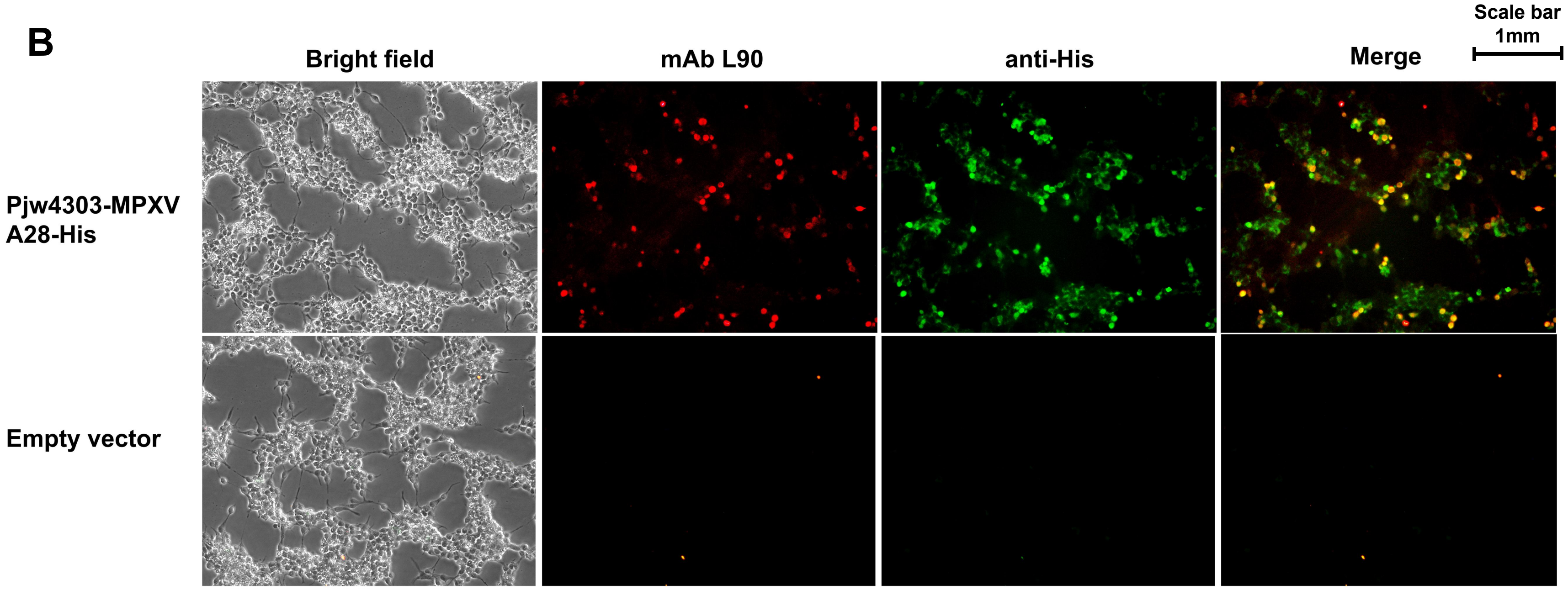


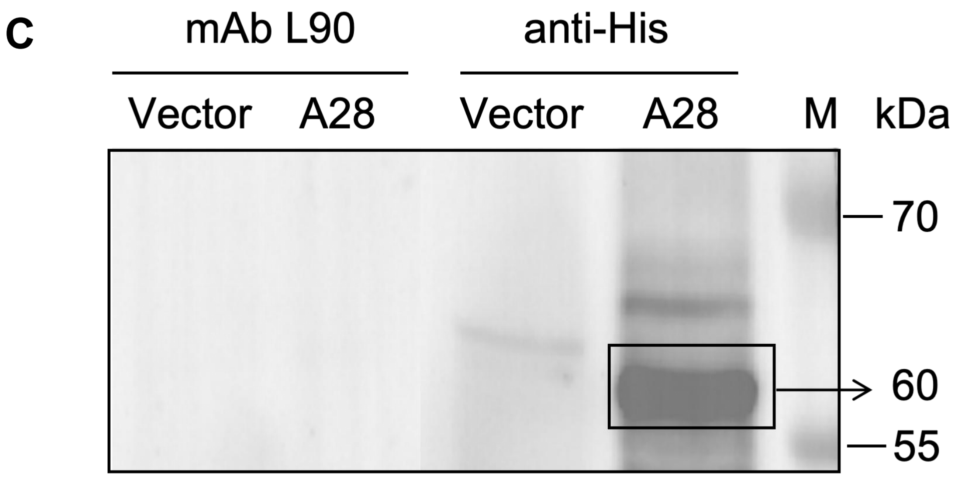

### Supplementary Figure 8

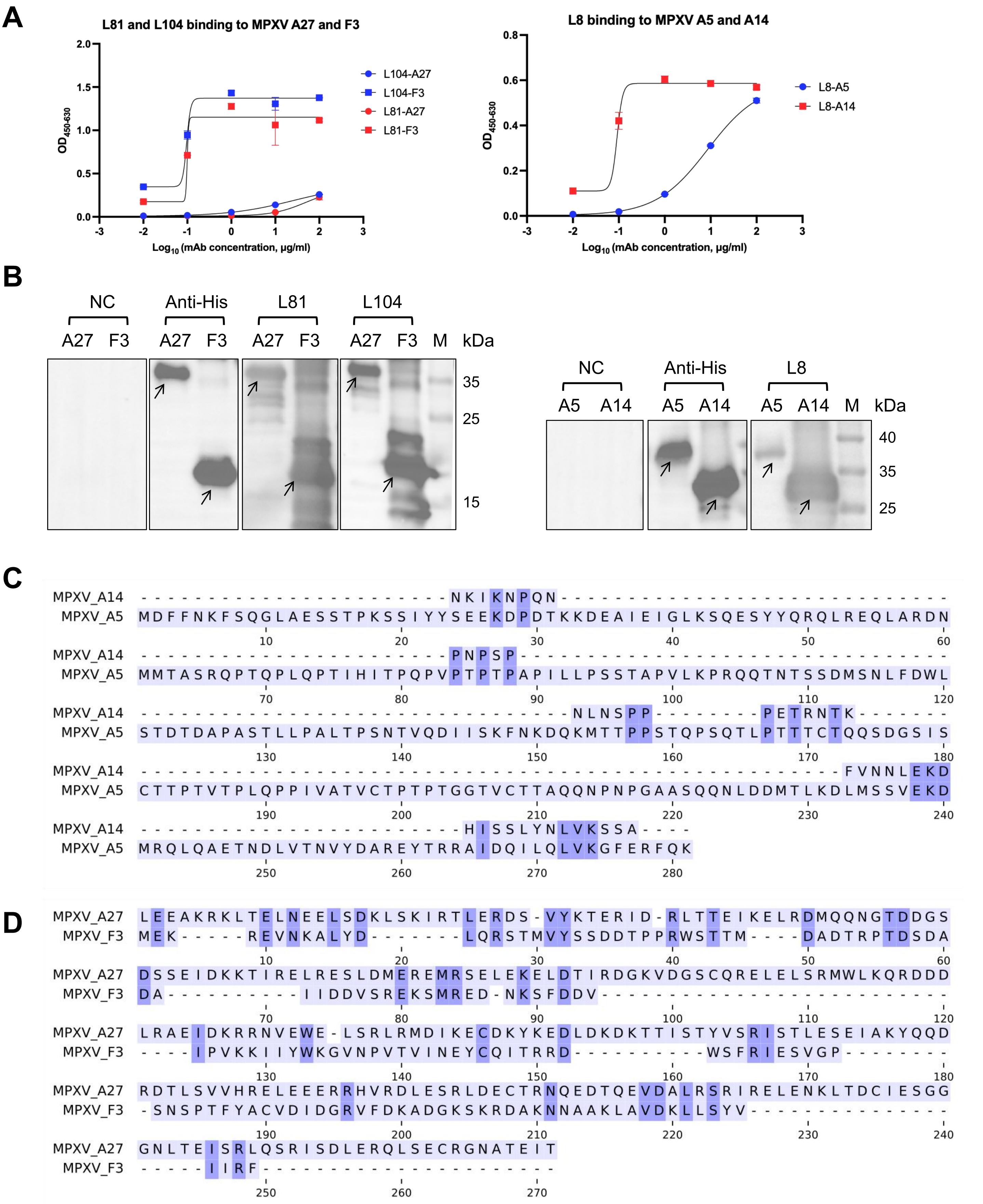

### Supplementary Figure 9

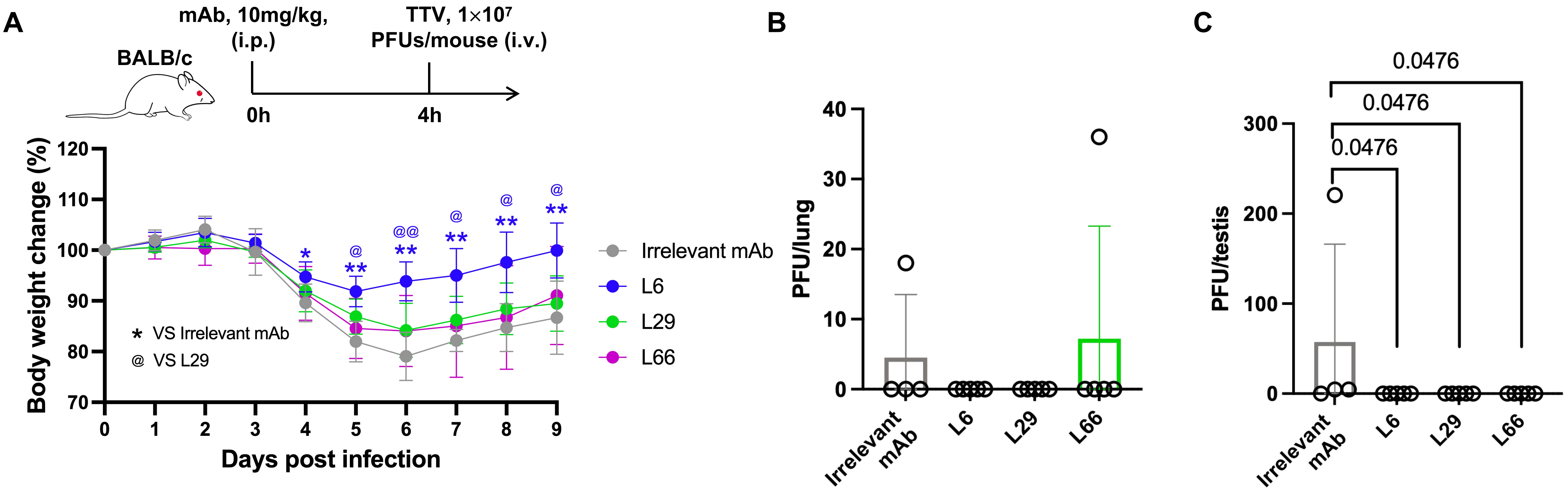

### Supplementary Figure 10

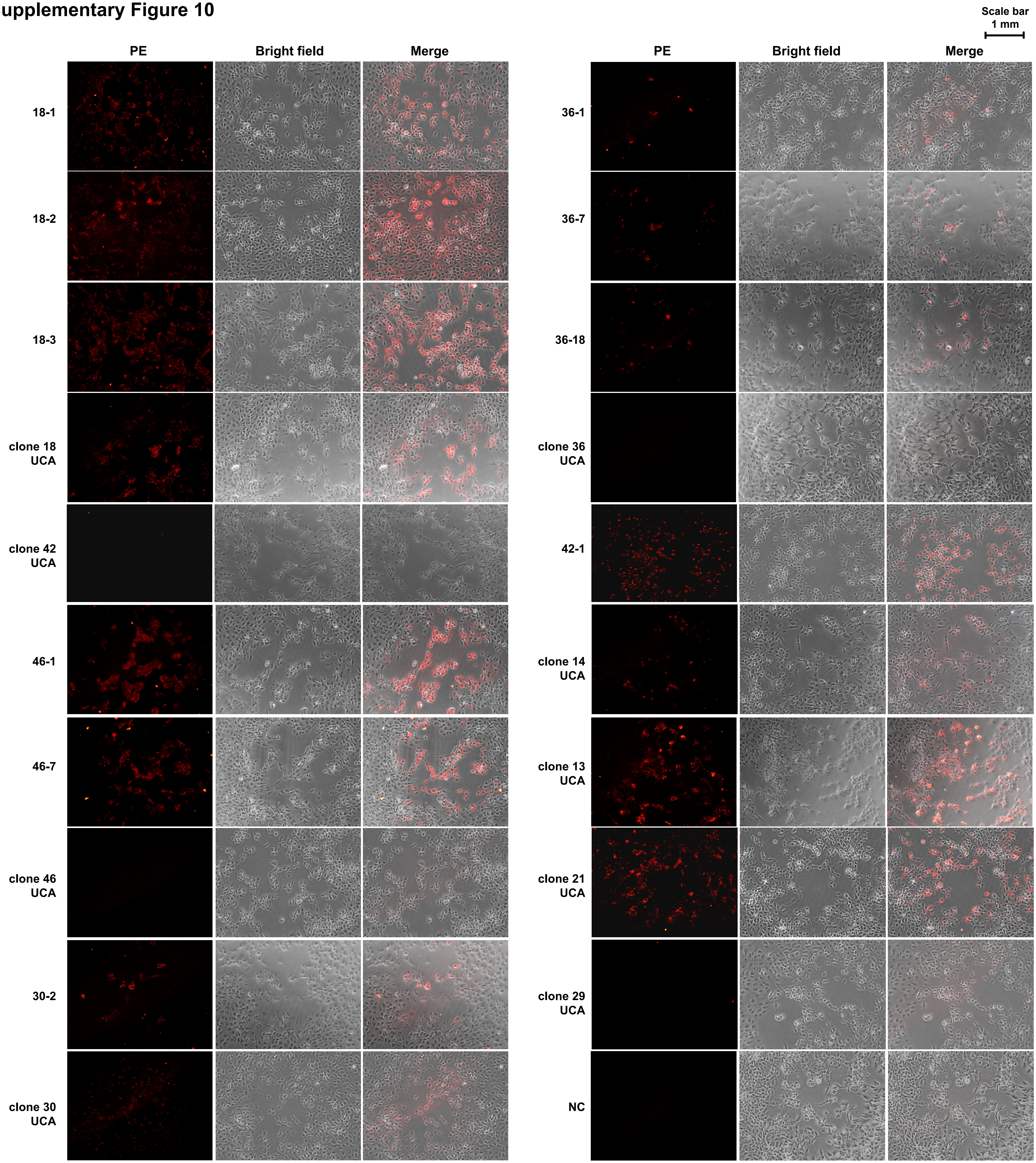

**Supplementary Figure 11**

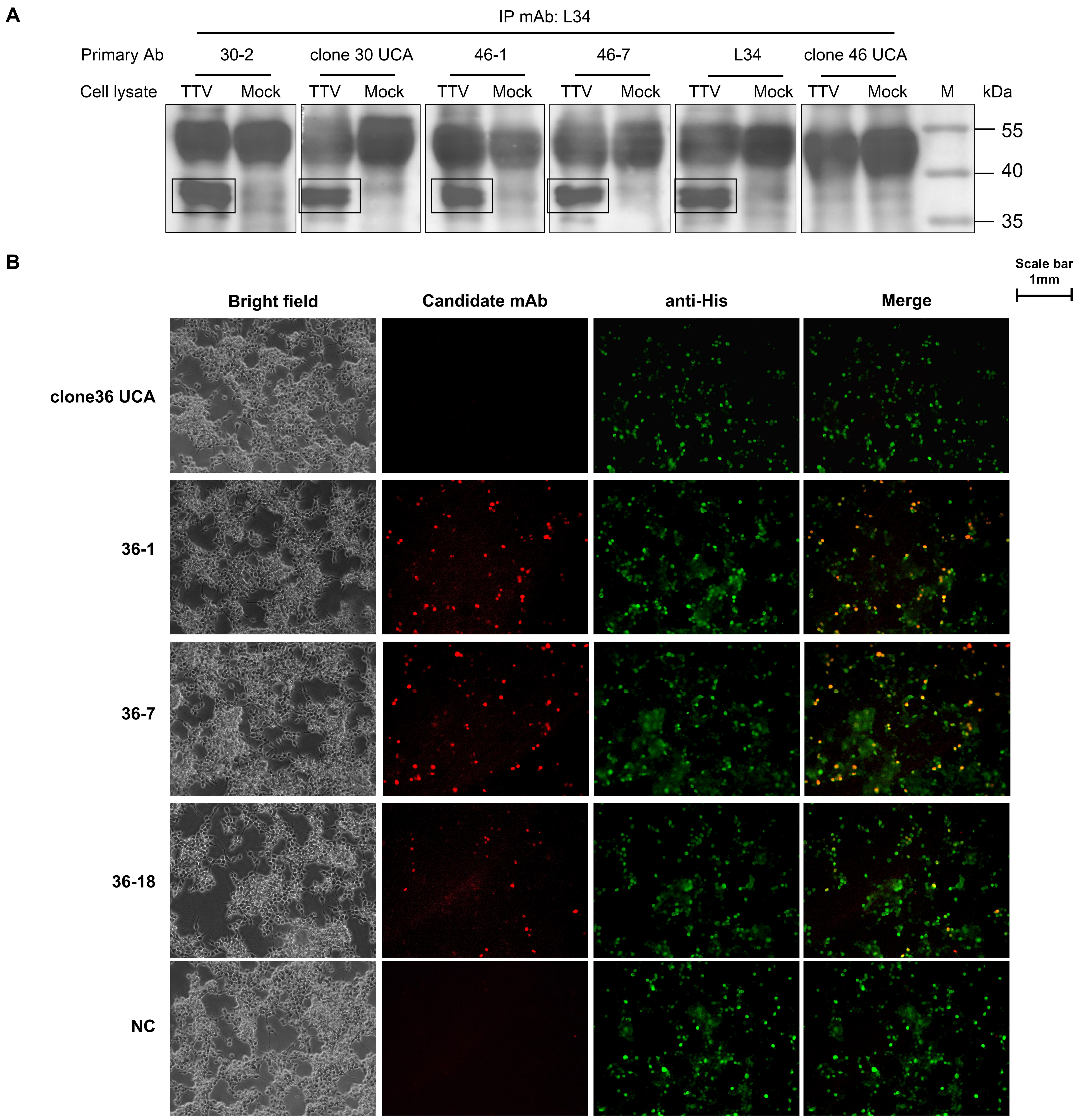

### Supplementary Figure 12

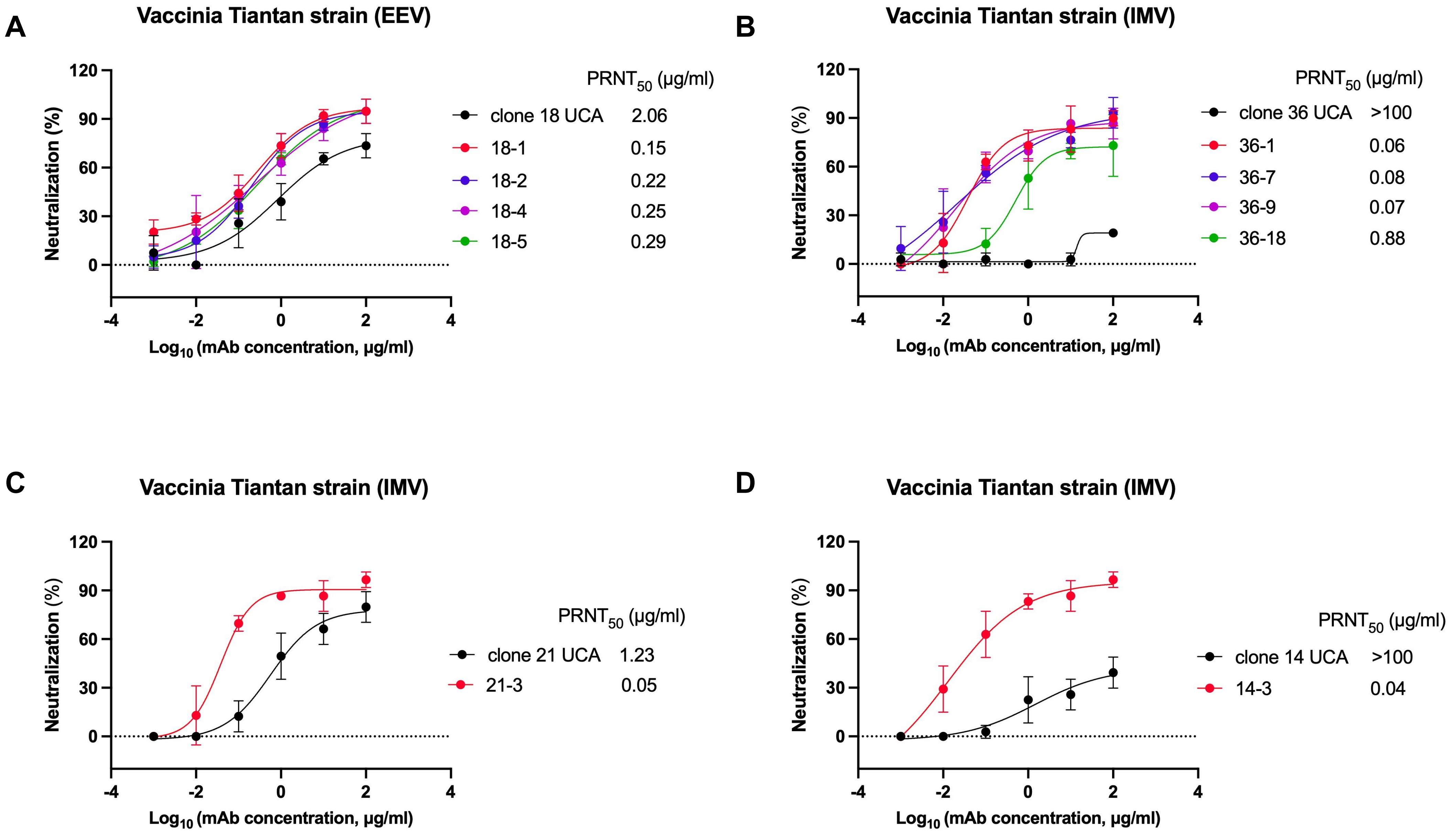

#### Supplementary Figure 13

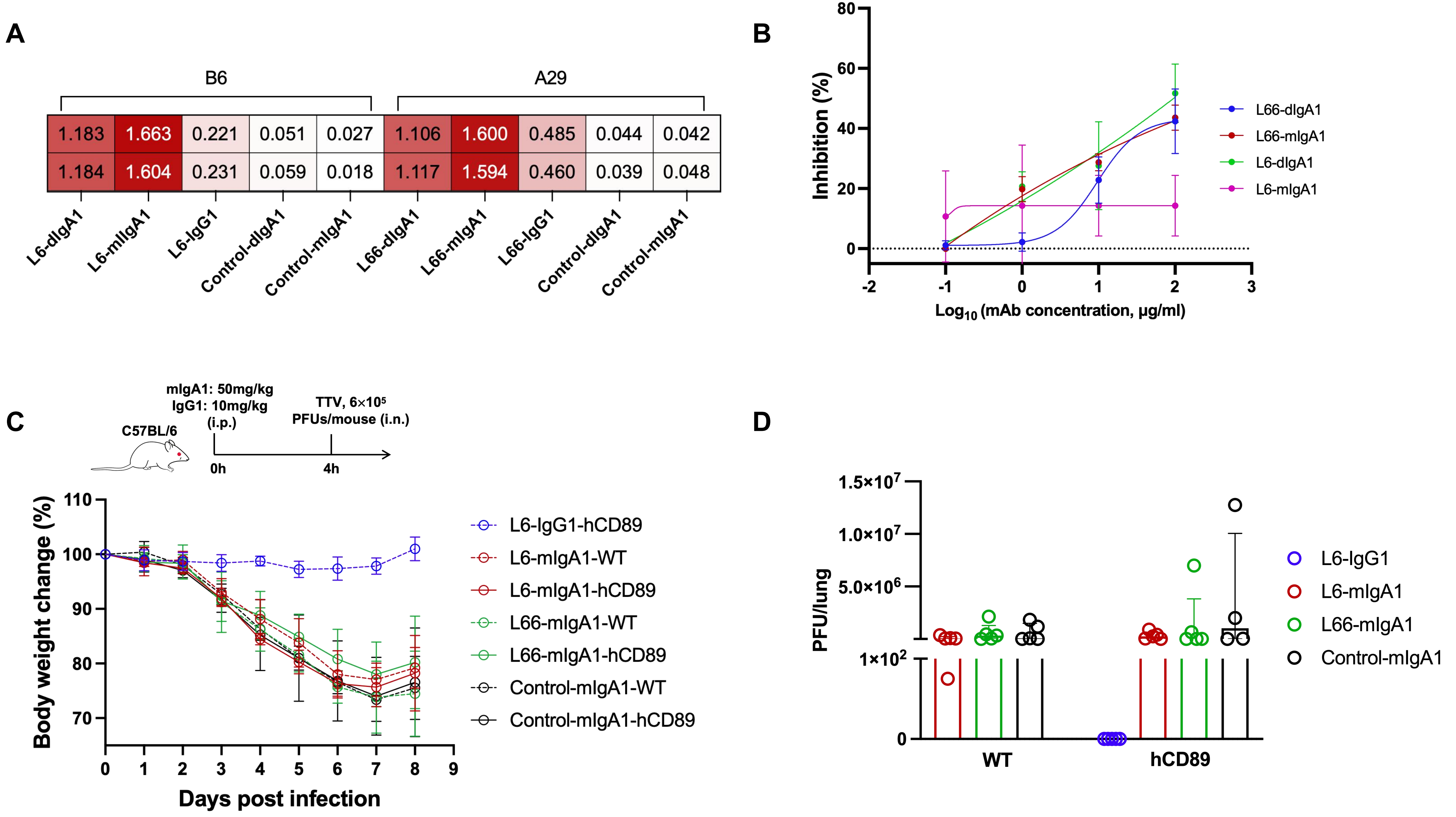
